# Impact of body composition on COVID-19 susceptibility and severity: a two-sample multivariable Mendelian randomization study

**DOI:** 10.1101/2020.07.14.20153825

**Authors:** Dennis Freuer, Jakob Linseisen, Christa Meisinger

**Affiliations:** Chair of Epidemiology at UNIKA-T Augsburg, Ludwig-Maximilians-Universität München, 86156 Augsburg, 80539 München, Germany; Independent Research Group Clinical Epidemiology, Helmholtz Center Munich, German Research Centre for Environmental Health, 85764 Neuherberg, Germany

**Keywords:** obesity, body fat distribution, body composition, COVID-19, SARS-CoV-2, Mendelian randomization

## Abstract

**Objectives:** Recent studies suggested obesity to be a possible risk factor for COVID-19 disease in the wake of the coronavirus (SARS-CoV-2) infection. However, the causality and especially the role of body fat distribution in this context is still unclear. Thus, using a univariable as well as multivariable two-sample Mendelian randomization (MR) approach, we investigated for the first time the causal impact of body composition on the susceptibility and severity of COVID-19.

**Methods:** As indicators of overall and abdominal obesity we considered the measures body mass index (BMI), waist circumference (WC), and trunk fat ratio (TFR). Summary statistics of genome-wide association studies (GWASs) for these body composition measures were drawn from the GIANT consortium and UK Biobank, while for susceptibility and severity due to COVID-19 disease data from the COVID-19 Host Genetics Initiative was used. For the COVID-19 cohort neither age nor gender was available. Total and direct causal effect estimates were calculated using Single Nucleotide Polymorphisms (SNPs), sensitivity analyses were done applying several robust MR techniques and mediation effects of type 2 diabetes (T2D) and cardiovascular diseases (CVD) were investigated within multivariable MR analyses.

**Results:** Genetically predicted BMI was strongly associated with both, susceptibility (OR=1.31 per 1 SD increase; 95% CI: 1.15–1.50; P-value=7.3·10^−5^) and hospitalization (OR=1.62 per 1 SD increase; 95% CI: 1.33–1.99; P-value=2.8·10^−6^) even after adjustment for genetically predicted visceral obesity traits. These associations were neither mediated substantially by T2D nor by CVD. Finally, total but not direct effects of visceral body fat on outcomes could be detected.

**Conclusions:** This study provides strong evidence for a causal impact of overall obesity on the susceptibility and severity of COVID-19 disease. The impact of abdominal obesity was weaker and disappeared after adjustment for BMI. Therefore, obese people should be regarded as a high-risk group. Future research is necessary to investigate the underlying mechanisms linking obesity with COVID-19.

## 1. INTRODUCTION

The global COVID-19 pandemic due to an outbreak of a coronavirus infection (SARS-CoV-2) causes serious conditions such as respiratory failure, pneumonia, and is associated with a high number of deaths[1]. Therefore, it is essential to identify risk factors associated with a higher susceptibility to COVID-19 or a more severe course of the disease and subsequently to identify high risk groups that require special protection[2]. Prior observational studies and meta-analyses found that SARS-CoV-2-patients with chronic diseases such as diabetes, cardiovascular diseases, chronic kidney disease, and respiratory diseases might be at increased risk of disease severity and mortality[3, 4]. Recent observational studies and systematic reviews and meta-analyses reported that also obese people may be vulnerable to a more severe COVID-19 disease course[5-8]. However, results were inconsistent on the association between BMI and COVID-19[9-12]. Using a Mendelian randomization (MR) approach, for very severe cases of COVID-19 patients with respiratory failure, BMI was already shown to have a causal impact[13, 14]. Another MR-study suggested genetically predicted BMI as causal risk factor for susceptibility and severity of COVID-19[15]. However, so far, research on the causal impact of body fat distribution on the susceptibility and severity of a COVID-19 disease is missing.

MR studies use genetic variants reliably related to a modifiable risk factor to obtain evidence regarding the causal influence of the risk factor. Thus, it is possible to minimize confounding and preclude reverse causation because variants are randomly allocated from parents to offspring at conception. In this study we investigated for the first time total as well as direct causal effects of an increase in BMI, waist circumference (WC), and trunk fat ratio (TFR) on the risk of infection and severe course of COVID-19 disease. We disentangled the suggested effects of overall and visceral body fat using refined statistical methods.

## 2. METHODS

### 2.1. Study design

In a two-sample MR approach genetic variants were used to assess causal effects of obesity considering also body fat distribution as risk factor on COVID-19 susceptibility and hospitalization due to an infection with the SARS-CoV-2 virus. Regarding the random allocation of single nucleotide polymorphisms (SNPs) at the offspring (independent of any confounders like gender and age and therefore no need for adjustment), this kind of instrumental variable analysis mimics a randomized controlled trial. The genetic variant(s) which are used in MR studies has to fulfill three key assumptions: 1) the SNPs must be robustly associated with the risk factor; 2) the SNPs are not associated with any confounder of the risk factor-outcome association (horizontal pleiotropy); 3) the genetic variant(s) affect the outcome solely through the risk factor and not through another causal pathway. While the univariable MR offers the possibility to examine causality by using genetic variants as instruments, which are explicitly associated with one exposure, the multivariable Mendelian randomization (MVMR) allows investigating causality of SNPs that are associated with more than one exposure. In this way, it is possible to distinguish between total and direct causal effects. The direct effect is defined as the effect of an exposure on the outcome only via one path (direct) but not via any other path [Figure 1]. By contrast, the total effect is defined as the sum of all possible paths from the exposure on the outcome. Further details on the MR- and MVMR-design were described elsewhere[16-19].

**Figure 1.**
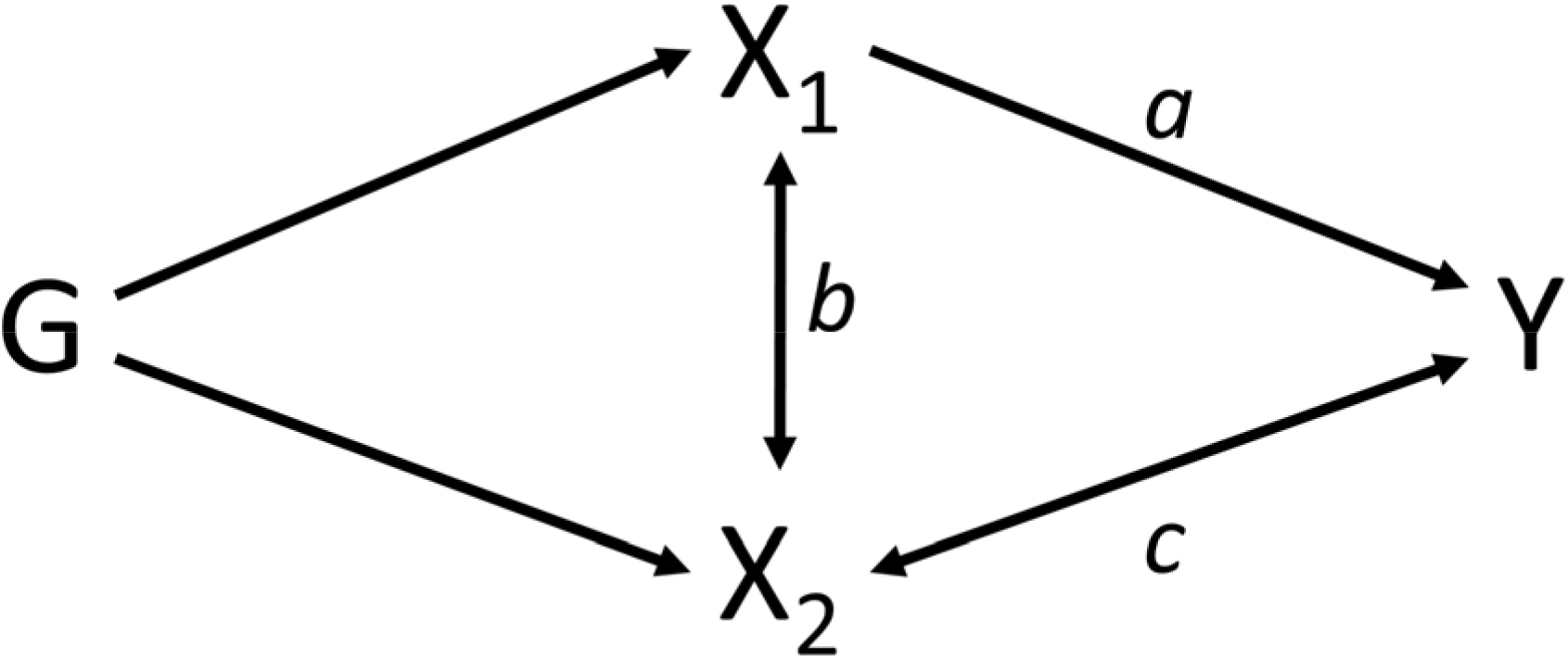
Simplified illustration showing the difference between a direct and total effect using two exposures in a multivariable Mendelian randomization setting. G represents a set of valid genetic instruments of two exposures X_1_ and/or X_2_. Bi-directional arrows represent possible violations of the IV assumptions induced by X_2_. The direct effect of exposure X_1_ on outcome Y is illustrated by the path a between X_1_ and Y. By contrast, the total effect of X_1_ is defined as the sum of all paths from X_1_ on Y (a, b and c).

### 2.2. Obesity measurements and data availability

To quantify obesity and to assess body fat distribution we used three different body composition measures, namely BMI as indicator for overall obesity, and WC and TFR as measures for visceral adiposity. TFR, defined as proportion of body fat in the trunk was calculated by dividing the fat mass of the trunk by total body fat mass that were determined by segmental bioelectrical impedance analysis[20]. For BMI, the summary statistics of a genome-wide association study (GWAS) conducted by Pulit et al.[21] including 694,649 participants from the GIANT consortium and UK Biobank [Table 1] were considered. To account for abdominal obesity we considered both the WC-GWAS of Shungin et al.[22] based on 232,101 observations from the GIANT consortium and the TFR-GWAS of Rask-Andersen et al.[20] with 362,499 subjects from the UK Biobank. The use of both datasets allowed us to replicate and verify the results and thus strengthen evidence. Furthermore, for mediation analyses the latest GWAS for type 2 diabetes (T2D) of the MRC IEU OpenGWAS Project[23] with 12,375 cases and 82,665 controls of the FinnGen Biobank and for cardiovascular disease the summary level data of Schunkert et al.[24] with 22,233 cases and 64,762 controls from the CARDIoGRAM consortium were used. Both mediation GWASs has the advantage that there is no sample overlap with the selected datasets of metabolic traits what is important to avoid issues regarding SNP-covariance estimation within mediation analysis[25]. Finally, in additional analyses we used the GWAS of Berndt et al.[26] with 98,697 participants from the GIANT consortium that considered distinct BMI categories subdivided in normal weight (BMI < 25 kg/m^2^), overweight (BMI ≥ 25 kg/m^2^), and obesity classes 1 – 3 with thresholds 30, 35, and 40, respectively. All datasets include observations in men and women of European ancestry. In all original studies, ethical approval had been obtained.

**Table 1.** Characteristics of the Single Nucleotide Polymorphisms (SNPs) used as instrumental variables for body mass index (BMI), waist circumference (WC), and trunk fat ratio (TFR) in the two-sample Mendelian randomization analyses

|  | BMI | WC | TFR |
| --- | --- | --- | --- |
| Sample size | 694,649 | 232 101 | 362,499 |
| Consortium | GIANT, UK Biobank | GIANT | UK Biobank |
| Number of instrumental SNPs<sup>a</sup> | 524 | 42 | 33 |
| Explained variance | 5.51 % | 1.09 % | 1.25 % |
| F-statistic (mean, range) | 72.29 (28.44 – 1,270.71) | 55.26 (29.34 – 447.02) | 41.69 (30.26 – 124.77) |
| Reference | Pulit et al, 2018 | Shungin et al, 2015 | Rask-Andersen et al, 2019 |

<sup>a</sup> extraction of SNPs based on the threshold $\alpha = 5 \cdot 10^{-8}$ and clumping cutoff $r^2 = 0.001$ .

### 2.3. Selection of genetic instrumental variables for body composition measures

As independent instruments we considered SNPs that were associated with the appropriate exposure at the genome-wide significance threshold P < 5·10^−8^ and were not in linkage disequilibrium (LD) using a clumping procedure with the cut-off r^2^ = 0.001. Therefore, 524 BMI-related, 42 WC-related and 33 TFR-related independent SNPs were considered for univariable MR analyses after removing SNPs with an imputation score ≤ 0.8 and palindromic SNPs with intermediate allele frequencies [Tables A.1 – A.3]. These SNPs explained overall 5.51 %, 1.09 %, and 1.25 % of the variance in BMI, WC, and TFR, respectively [Table 1]. Mean F-statistics, as measure for instrument strength, ranged from 41.69 (TFR) to 72.29 (BMI) indicating strong instruments and therefore low predisposition for weak instruments bias. Even the lowest F-statistic F=28.44 in BMI was above the suggested threshold for sufficient instrument strength of F=10.

### 2.4. GWAS summary statistics for COVID-19 disease

Genetic associations with COVID-19 susceptibility and hospitalization were acquired from the growing COVID-19 Host Genetics Initiative[27], which provides publicly accessible summary statistics of GWAS in relation to several COVID-19 outcomes from different studies (e.g. UK Biobank, FinnGen). In the third release from June 29, 2020 in total 6696 positively tested COVID-19 cases (vs. 1073072 controls) and 3199 hospitalized COVID-19 patients due to severe symptoms (vs. 897488 controls) were available. However, neither age nor gender was reported for the cohort. Further information and new releases can be taken from the COVID-19 Host Genetics Initiative homepage[27].

### 2.5. Statistical power

The a priori statistical power for the binary traits susceptibility and hospitalization due to COVID-19 disease was calculated according to Burgess et al.[28]. Given a type I error of 5 %, the power for susceptibility was higher than for hospitalization and higher for BMI than for WC or TFR regarding the explained variance in exposures as well as the number of cases/controls in the outcomes [Table A.4]. Therefore, our analyses were sufficiently powered (i.e. > 80 %) when the true OR per one standard deviation of the respective exposure was ≥ 1.4 for COVID-19 susceptibility and ≥ 1.5 for hospitalization due to COVID-19 in genetically instrumented BMI, WC and TFR.

### 2.6. Statistical analyses

Causal estimates of the relationships between body composition measures and Covid-19 susceptibility as well as severity were calculated, applying an inverse-variance weighted (IVW) meta-analysis using modified second order weights within the radial regression framework. This approach provides the highest statistical power if the key assumptions of the MR (described in the study design) are met. To assess and validate these assumptions we performed a series of sensitivity analyses that consider different patterns of violations. Among others, the MR-PRESSO (Pleiotropy RESidual Sum and Outlier) method was used for two issues: the global test (based on observed residual sum of squares) detected horizontal pleiotropy and the outlier test identified potential outlier SNPs at a threshold of 0.05 [Table A.5].

Under the Instrument Strength Independent of Direct Effect (InSIDE) assumption, the radial MR-Egger intercept test was applied for assessing directional pleiotropy [Table A.5]. Substantial heterogeneity within the IVW and MR-Egger methods was quantified and tested using Cochran’s as well as Rücker’s Q statistics [Table A.6]. Furthermore, we investigated influential SNPs based on the respective Q statistic and several plots (e.g. radial, funnel, leave-one-out, and SNP-exposure-outcome association plots). If necessary, outliers were excluded as a part of a sensitivity analysis within an iterative approach [Table A.7].

To assess consistency of causal estimates in case of horizontal pleiotropy as well as outlier occurrence, we additionally performed four types of robust estimation methods. The first was the MR-Egger regression, which provides under the InSIDE assumption a consistent estimator, even if there is directional pleiotropy. The second was the weighted median approach that requires at least 50 % of the genetic variants to be valid instruments. The third was the weighted mode method, which is consistent even if less than 50 % of the genetic variants are valid. Fourth, we conducted a many weak instruments analysis using the RAPS (Robust Adjusted Profile Score) method and controlled for systematic pleiotropy in form of overdispersion, if necessary.

The used body composition measures for overall and visceral obesity are not independent from each other. Furthermore, the causal effect estimates of these traits on the susceptibility and severity of COVID-19 disease may be mediated through obesity-related diseases. Therefore, to calculate the direct effects of each body composition measure and to investigate possible mediation mechanisms of type 2 diabetes and cardiovascular diseases (CVD), we additionally performed MVMR analyses. In the main analysis we performed the robust IVW method with multiplicative random effects, where we mutually adjusted BMI as measure for overall body fat for the association of variants with genetically predicted levels of one of the visceral obesity traits (WC, TFR) or comorbidity (type 2 diabetes, CVD). Within sensitivity analyses the MR-Egger with multiplicative random effects and the Median approach were performed. In addition to this, we determined Q-minimized point estimates that take into account weak instruments and substantial heterogeneity using estimated phenotypic correlations. To assess substantial heterogeneity and directional pleiotropy, we calculated the global Q-statistics and applied the MR-Egger intercept test to all models, respectively [Tables A.8 -A.11].

To verify the results regarding overall fat content from univariable MR and make statements about obesity as a disease compared to non-obese subjects, additional analyses were performed using an independent GWAS with categorized BMI (five BMI groups) [26].

After application of the Bonferroni correction to account for multiple testing issues, P-values with P < 0.008 were considered statistically significant. All reported ORs were expressed per one standard deviation increment of each body composition measure. Significance thresholds were set to *α* = 0.01 for testing particular Q statistics and *α* = 0.05 for the PRESSO global test. Analyses were performed using primarily the TwoSampleMR (version 0.5.4), MendelianRandomization (version 0.5.0), MVMR (version 0.2) and MRPRESSO (version 1.0) packages of the statistical Software R (version: 4.0.0).

## 3. RESULTS

### 3.1. Total effects

Genetically predicted BMI was strongly positively associated with both, COVID-19 susceptibility (OR = 1.31; 95 % CI: 1.15 – 1.50; P-value = 7.3·10^−5^) and hospitalization (OR = 1.62; 95 % CI: 1.33 – 1.99; P-value = 2.8·10^−6^) due to severe symptoms of COVID-19 [Tables 2-3]. While genetically predicted TFR showed a strong relationship with disease susceptibility (OR = 1.42; 95 % CI: 1.13 – 1.78; P-value = 0.003), the association with severity was weaker (OR = 1.56; 95 % CI: 1.01 – 2.40; P-value = 0.044). Beyond that, there was evidence of a positive association between WC and COVID-19 susceptibility (OR = 1.38; 95 % CI: 1.07 – 1.78; P-value = 0.015) but not with hospitalization due to COVID-19 (OR = 1.47; 95 % CI: 0.97 – 2.23; P-value = 0.069).

**Table 2.** Univariable Mendelian randomization estimates (total effects) for the association between body mass index (BMI), waist circumference (WC), and trunk fat ratio (TFR) related Single Nucleotide Polymorphisms (SNPs) and COVID-19 susceptibility

| Method | $n_{SNP}$ | OR | 95 % CI | P | Exposure |
| --- | --- | --- | --- | --- | --- |
| Radial IVW (Mod.2nd) | 519 | 1.312 | (1.147 - 1.500) | $7.3 \cdot 10^{-5}$ | BMI |
| Radial IVW (exact fixed effects) | 519 | 1.317 | (1.148 - 1.510) | $8.1 \cdot 10^{-5}$ | BMI |
| Radial IVW (exact random effects) | 519 | 1.317 | (1.153 - 1.504) | $5.5 \cdot 10^{-5}$ | BMI |
| IVW (mult. random effects) | 519 | 1.312 | (1.147 - 1.500) | $7.3 \cdot 10^{-5}$ | BMI |
| RAPS | 519 | 1.317 | (1.146 - 1.513) | $1.0 \cdot 10^{-4}$ | BMI |
| PRESSO | 519 | 1.312 | (1.147 - 1.500) | $8.3 \cdot 10^{-5}$ | BMI |
| Weighted median | 519 | 1.333 | (1.077 - 1.650) | 0.008 | BMI |
| Weighted mode | 519 | 1.414 | (0.887 - 2.255) | 0.146 | BMI |
| MR-Egger | 519 | 1.297 | (0.884 - 1.905) | 0.184 | BMI |
| Radial IVW (Mod.2nd) | 41 | 1.377 | (1.065 - 1.780) | 0.015 | WC |
| Radial IVW (exact fixed effects) | 41 | 1.384 | (1.040 - 1.843) | 0.026 | WC |
| Radial IVW (exact random effects) | 41 | 1.384 | (1.089 - 1.760) | 0.011 | WC |
| IVW (mult. random effects) | 41 | 1.377 | (1.065 - 1.780) | 0.015 | WC |
| RAPS | 41 | 1.384 | (1.033 - 1.855) | 0.029 | WC |
| PRESSO | 41 | 1.377 | (1.065 - 1.780) | 0.019 | WC |
| Weighted median | 41 | 1.362 | (0.904 - 2.053) | 0.139 | WC |
| Weighted mode | 41 | 1.296 | (0.670 - 2.507) | 0.445 | WC |
| MR-Egger | 41 | 1.817 | (0.654 - 5.050) | 0.259 | WC |
| Radial IVW (Mod.2nd) | 32 | 1.417 | (1.126 - 1.783) | 0.003 | TFR |
| Radial IVW (exact fixed effects) | 32 | 1.424 | (1.069 - 1.897) | 0.016 | TFR |
| Radial IVW (exact random effects) | 32 | 1.424 | (1.136 - 1.785) | 0.004 | TFR |
| IVW (mult. random effects) | 32 | 1.417 | (1.126 - 1.783) | 0.003 | TFR |
| RAPS | 32 | 1.424 | (1.061 - 1.913) | 0.019 | TFR |
| PRESSO | 32 | 1.417 | (1.126 - 1.783) | 0.006 | TFR |
| Weighted median | 32 | 1.411 | (0.944 - 2.109) | 0.093 | TFR |
| Weighted mode | 32 | 1.526 | (0.766 - 3.041) | 0.238 | TFR |
| MR-Egger | 32 | 1.746 | (0.564 - 5.405) | 0.341 | TFR |

Abbreviations: OR, odds ratio; CI, confidence interval; IVW (Mod.2nd) inverse-variance weighted model with modified 2<sup>nd</sup> order weights.

**Table 3.** Univariable Mendelian randomization estimates (total effects) for the association between body mass index (BMI), waist circumference (WC), and trunk fat ratio (TFR) related Single Nucleotide Polymorphisms (SNPs) and hospitalization due to severity of COVID-19 disease

| Method | $n_{SNP}$ | OR | 95 % CI | P | Exposure |
| --- | --- | --- | --- | --- | --- |
| Radial IVW (Mod.2nd) | 518 | 1.623 | (1.325 - 1.989) | $2.8 \cdot 10^{-6}$ | BMI |
| Radial IVW (exact fixed effects) | 518 | 1.635 | (1.336 - 2.002) | $1.8 \cdot 10^{-6}$ | BMI |
| Radial IVW (exact random effects) | 518 | 1.635 | (1.340 - 1.996) | $1.7 \cdot 10^{-6}$ | BMI |
| IVW (mult. random effects) | 518 | 1.623 | (1.325 - 1.989) | $2.8 \cdot 10^{-6}$ | BMI |
| RAPS | 518 | 1.635 | (1.332 - 2.008) | $2.6 \cdot 10^{-6}$ | BMI |
| PRESSO | 518 | 1.623 | (1.325 - 1.989) | $3.6 \cdot 10^{-6}$ | BMI |
| Weighted median | 518 | 1.544 | (1.103 - 2.161) | 0.011 | BMI |
| Weighted mode | 518 | 1.271 | (0.629 - 2.571) | 0.504 | BMI |
| MR-Egger | 518 | 1.270 | (0.721 - 2.237) | 0.407 | BMI |
| Radial IVW (Mod.2nd) | 41 | 1.472 | (0.971 - 2.232) | 0.069 | WC |
| Radial IVW (exact fixed effects) | 41 | 1.483 | (0.971 - 2.265) | 0.068 | WC |
| Radial IVW (exact random effects) | 41 | 1.483 | (0.992 - 2.217) | 0.062 | WC |
| IVW (mult. random effects) | 41 | 1.472 | (0.971 - 2.232) | 0.069 | WC |
| RAPS | 41 | 1.483 | (0.962 - 2.285) | 0.074 | WC |
| PRESSO | 41 | 1.472 | (0.971 - 2.232) | 0.076 | WC |
| Weighted median | 41 | 1.441 | (0.788 - 2.633) | 0.235 | WC |
| Weighted mode | 41 | 1.240 | (0.442 - 3.477) | 0.685 | WC |
| MR-Egger | 41 | 1.640 | (0.365 - 7.360) | 0.523 | WC |
| Radial IVW (Mod.2nd) | 33 | 1.560 | (1.013 - 2.404) | 0.044 | TFR |
| Radial IVW (exact fixed effects) | 33 | 1.577 | (1.036 - 2.402) | 0.034 | TFR |
| Radial IVW (exact random effects) | 33 | 1.576 | (1.028 - 2.418) | 0.045 | TFR |
| IVW (mult. random effects) | 33 | 1.560 | (1.013 - 2.404) | 0.044 | TFR |
| RAPS | 33 | 1.577 | (1.025 - 2.427) | 0.038 | TFR |
| PRESSO | 33 | 1.560 | (1.013 - 2.404) | 0.052 | TFR |
| Weighted median | 33 | 1.643 | (0.906 - 2.979) | 0.102 | TFR |
| Weighted mode | 33 | 1.700 | (0.519 - 5.566) | 0.387 | TFR |
| MR-Egger | 33 | 3.254 | (0.565 - 18.751) | 0.196 | TFR |

Abbreviations: OR, odds ratio; CI, confidence interval; IVW (Mod.2nd) inverse-variance weighted model with modified 2<sup>nd</sup> order weights.

We conducted a series of sensitivity analyses to assess the robustness of the results. Neither the MR-Egger intercept test nor the MR-PRESSO global test nor the calculated Q-statistics provided evidence for any pleiotropy [Tables A.5 - A.6]. Outlier exclusion [Table A.7] had no substantial effect on the appropriate estimates [Figure 2]. Therefore, causal estimates were obtained by the radial IVW model using modified second order weights. All robust approaches within sensitivity analyses led to consistent results and supported the findings revealed by the radial IVW models [Tables 2-3].

**Figure 2.**
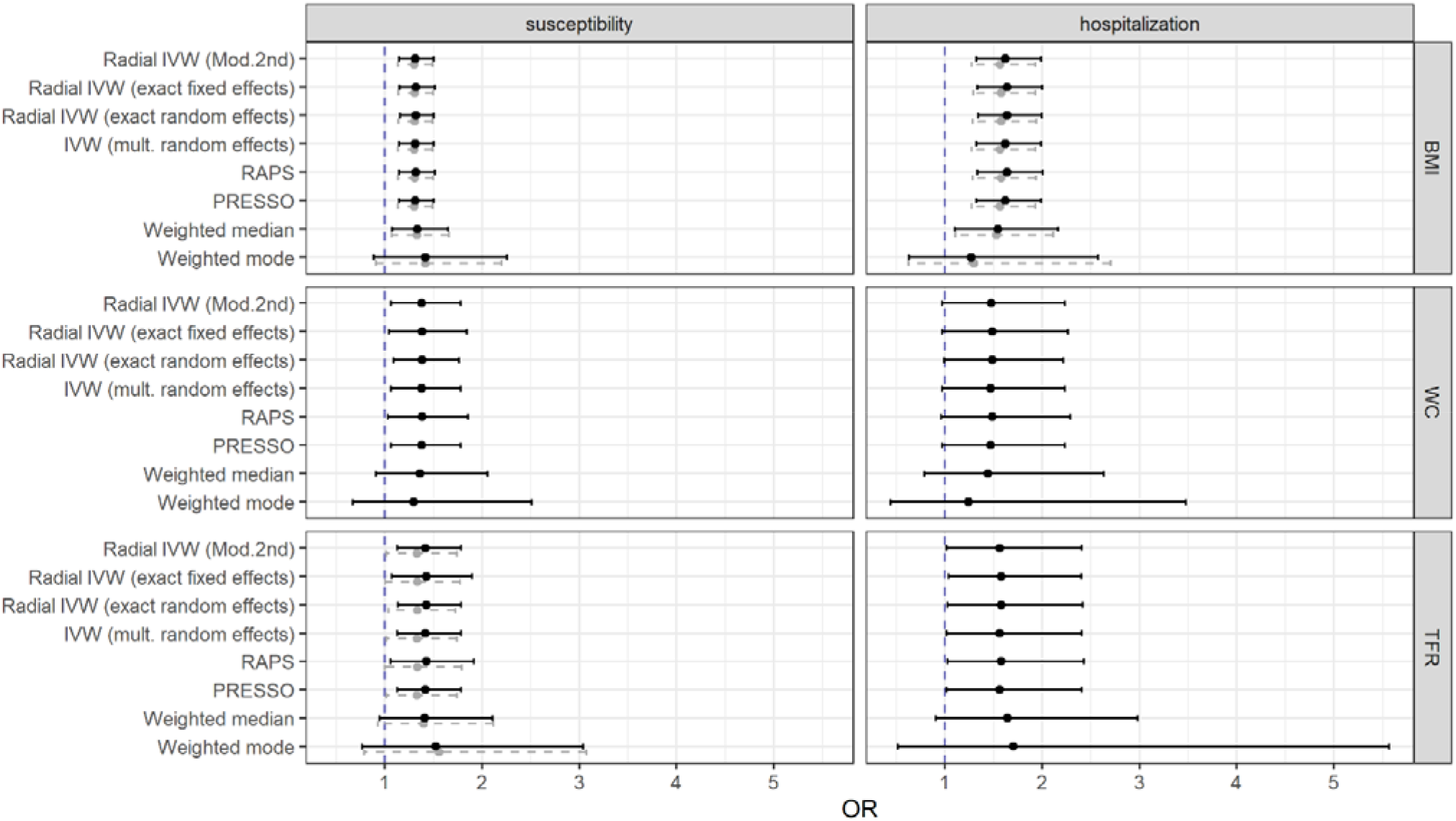
Causal total effect estimates (odds ratios and 95 % confidence intervals) from the univariable Mendelian randomization analyses of body mass index (BMI), waist circumference (WC), and trunk fat ratio (TFR) with COVID-19 susceptibility and hospitalization. Grey points with dashed confidence intervals correspond to estimates biased regarding influential SNPs [Table A.7]. For reasons of clarity MR-Egger estimates were omitted regarding wide confidence intervals. Abbrevations: IVW (Mod.2nd) inverse-variance weighted model with modified 2^nd^ order weights.

### 3.2. Direct effects

After mutual adjustment for WC or TFR within the MVMR setting, genetically predicted BMI was still positively associated with both, COVID-19 susceptibility and severity with odds ratios between OR_TFR adj_ = 1.29 (95 % CI: 1.07 – 1.54; P-value = 0.007) and OR_WC adj_ = 2.13 (95 % CI: 1.17 – 3.89; P-value = 0.014) [Figure 3]. Beyond that, the effect estimates of both visceral obesity traits lost their statistical significance after adjustment for BMI. The MVMR-Egger approach with multiplicative random effects as well as the Median method supported largely these findings [Fig. A.12]. However, the Egger intercept test in the model of BMI together with TFR on the hospitalization due to COVID-19 revealed directional pleiotropy (*β*_intercept_ = 0.01; 95 % CI: 0.00 – 0.02; P-value = 0.044) [Table A.8] and a non-significant MR-Egger estimator for BMI in a different direction (OR = 0.74; 95 % CI: 0.36 – 1.51; P-value = 0.408) [Fig. A.12]. Apart from this, there was no evidence for directional pleiotropy [Table A.8] or heterogeneity [Table A.9] in the multivariable models, what was supported by the point estimates of the multivariable Q-minimization approach, which is even consistent even with weak instruments [Figure 3].

**Figure 3.**
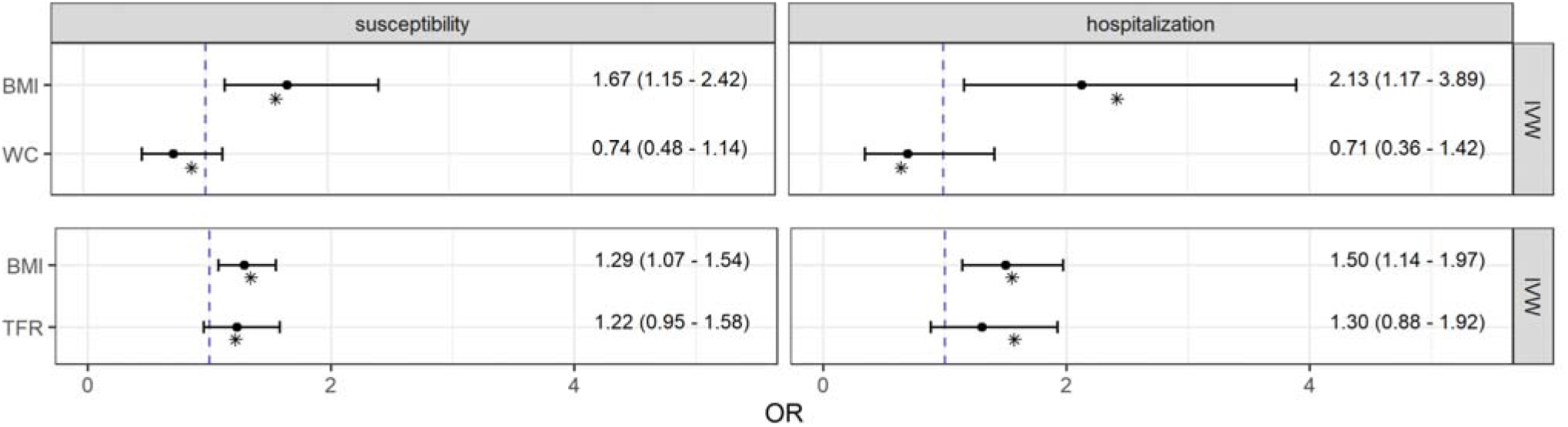
Causal direct effect estimates from pairwise multivariable Mendelian randomization analyses of body mass index (BMI), waist circumference (WC), and trunk fat ratio (TFR) with COVID-19 susceptibility as well as hospitalization. Odds ratios and 95 % confidence intervals were obtained from the robust inverse-variance weighted (IVW) method with multiplicative random effects. Point estimates shown as asterisks were obtained from the Q-minimization approach that account for weak instruments and substantial heterogeneity.

### 3.3. Mediation analysis

Neither type 2 diabetes nor CVD nor both together mediated the effects of body composition measures on the COVID-19 outcomes [Fig. A.13 - A.15]. Compared to the unadjusted OR the direct effect estimates of BMI on both susceptibility and severity due COVID-19 remained the same or increased slightly after adjustment [Figure 4]. In the joint models with both confounding variables the effect of BMI on the COVID-19 susceptibility was OR = 1.38 (95 % CI: 1.15 – 1.66; P-value = 4.7·10^−4^) and the effect on hospitalization was OR = 1.65 (95 % CI: 1.21 – 2.26; P-value = 0.002).

**Figure 4.**
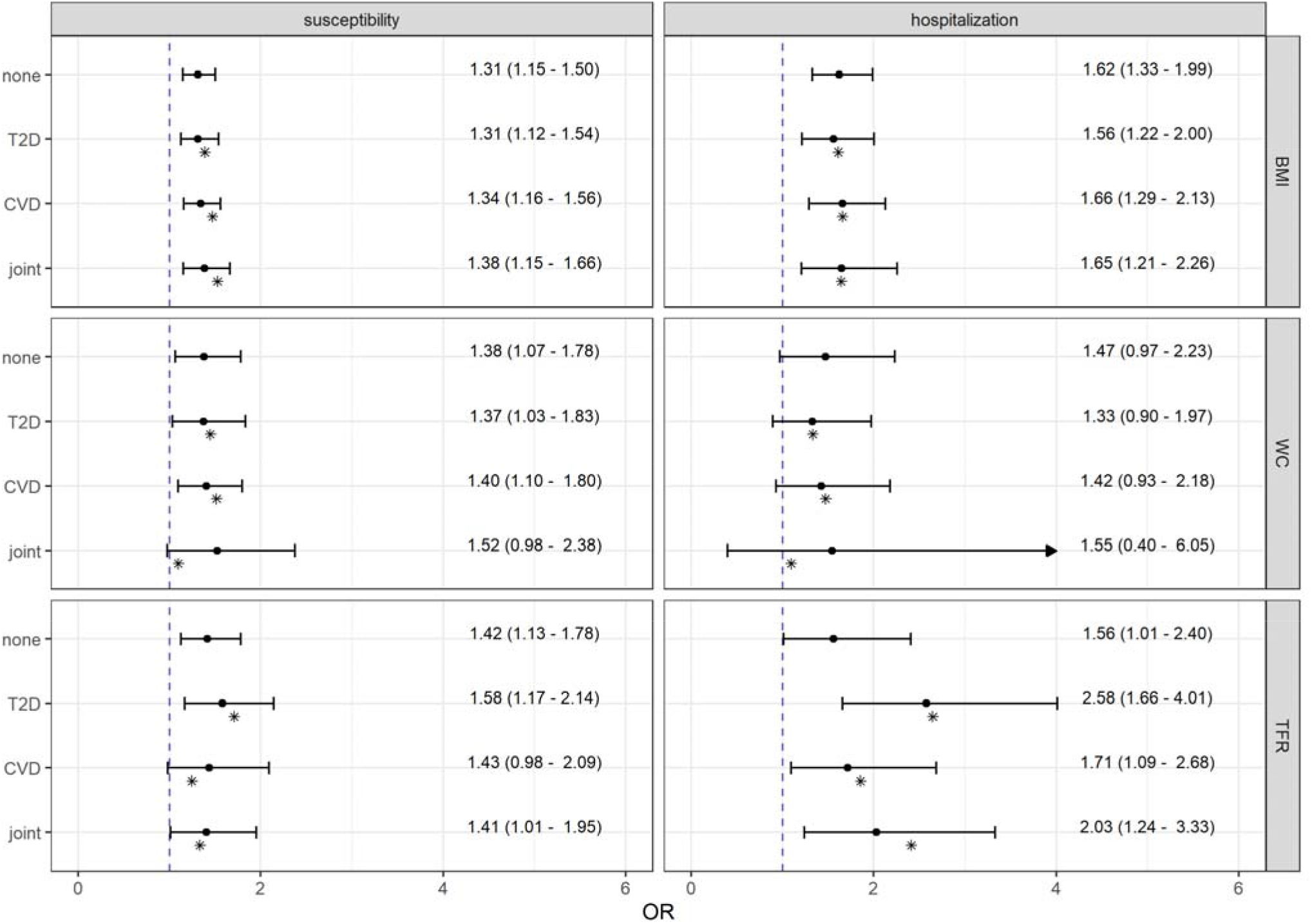
Total and direct effect estimates from Mendelian randomization mediation analyses of body composition measures, body mass index (BMI), waist circumference (WC), and trunk fat ratio (TFR), adjusted for type 2 diabetes (T2D) and/or cardiovascular diseases (CVD) on COVID-19 susceptibility as well as hospitalization. Odds ratios and 95 % confidence intervals were obtained from the robust inverse-variance weighted method with multiplicative random effects. Point estimates shown as asterisks were obtained from the Q-minimization approach that account for weak instruments and substantial heterogeneity.

Furthermore, except for the joint model, no noticeable impact of type 2 diabetes or CVD could be observed for the association between genetically predicted WC and susceptibility to COVID-19 with direct effects similar to the total effects.

The adjusted causal estimates for the impact of genetically predicted TFR on hospitalization due to COVID-19 ranging between OR_CVD adj_ = 1.71 (95 % CI: 1.09 – 2.68; P-value = 0.019) and OR_T2D adj_ = 2.58 (95 % CI: 1.66 – 4.01; P-value = 2.6·10^−5^) were stronger than the total effect estimates. This implies that TFR was neither attenuated by type 2 diabetes nor CVD nor by both together. However, the association of genetically predicted TFR with susceptibility to an infection with COVID-19 was affected slightly by CVD (OR_CVD adj_ = 1.43; 95 % CI: 0.98 – 2.09; P-value = 0.061), but neither by type 2 diabetes (OR_T2D adj_ = 1.58; 95 % CI: 1.17 – 2.14; P-value = 0.003) nor in the joint model (OR_joint_ = 1.41; 95 % CI: 1.01 – 1.95; P-value = 0.043).

All estimates replicated by MVMR-Egger as well as Median approaches in our sensitivity analyses supported unfailingly the presented findings obtained by the robust IVW method with multiplicative random effects [Fig. A.13 – A.15]. Furthermore, there was no evidence for directional pleiotropy [Table A.10]. However, although the calculated Q-statistics revealed substantial heterogeneity in the joint models and type 2 diabetes-adjusted models of BMI on susceptibility and hospitalization due to COVID-19 [Table A.11], the calculated point estimates from the multivariable Q-minimization approach confirmed the appropriate findings of our mediation analyses [Figure 4].

### 3.4. Additional analysis

Overweight and obese subjects showed an increased risk for susceptibility and severity of COVID-19 compared to controls with a BMI < 25 kg/m^2^ [Fig. A.16]. Despite the relatively small sample size and therefore higher uncertainty, causal estimates were throughout consistently positive and rose with increasing obesity up to an OR = 1.04 (95 % CI: 1.03 – 1.05; P-value = 2.5·10^−14^) for susceptibility and an OR = 1.17 (95 % CI: 1.14 – 1.19; P-value = 8.2·10^−41^) for hospitalization due to COVID-19 disease. As before, all estimates derived by the IVW approach with multiplicative second order weights as main method were confirmed within the scope of sensitivity analyses and supported therefore the results from the univariable MR main analyses.

## 4. DISCUSSION

The present study using genetic instruments for BMI, WC, and TFR from publicly available large-scale GWAS provides evidence for a causal role of general obesity expressed by an increasing BMI regarding COVID-19 susceptibility and disease severity. The association was strongly positive even after adjustment for genetically predicted visceral fat. Adjusting for the genetic effects of type 2 diabetes and CVD, the direct causal effects of BMI were not attenuated and thus, a mediating effect of these comorbidities seems unlikely. In view of the visceral obesity traits, especially the TFR had strong total effects on both outcomes in the univariable analyses. However, after adjustment for CVD the association with COVID-19 susceptibility was slightly attenuated.

### 4.1. Literature context

The present MR-analysis supports recently published observational studies, systematic reviews and meta-analyses assuming that obesity defined by BMI might be an independent risk factor for COVID-19[9-12, 29]. In addition, the results of this MR-analysis clearly indicated a causal association of BMI and COVID-19 susceptibility and severity independent of the comorbidities type 2 diabetes and CVD. This finding is in line with two other MR-studies investigating the among other risk factors the association between genetically predicted BMI and COVID-19 severity[13, 14]. One further prior MR-study on cardiometabolic risk factors associated with COVID-19 identified BMI as a causal risk factor for COVID-19 susceptibility and severity[15]. Contrary to our findings, in that investigation, the authors concluded that the association between BMI and COVID-19 illness might be mediated by type 2 diabetes. However, in contrast to the study of Leong et al. in our investigation we used the latest mediation GWASs with no sample overlap with the selected datasets of body composition measures. This approach circumvented issues with estimations of phenotypic covariances and therefore minimized bias within the mediation analyses[25].

So far, studies investigating the role of body fat distribution on the severity and susceptibility of COVID-19 are scarce. Few observational studies investigated the association between visceral fat and COVID-19 disease severity and complications and suggested that abdominal fat is related to disease severity[30-34]. For example, in their proof-of-concept study including 30 patients with COVID-19 Petersen et al. suggested that visceral fat and upper abdominal circumference specifically increased the likelihood of severe COVID-19[32]. Another study showed that visceral adiposity and high intramuscular fat deposition assessed by computed tomography scans were independently associated with critical COVID-19 illness, that is patients with acute respiratory distress syndrome or sepsis with acute organ dysfunction[33]. Our results show that, apart from BMI, body fat distribution, in particular visceral adiposity, plays no direct causal role regarding COVID-19 severity and susceptibility. Observational studies on correlations of visceral fat accumulation with COVID-19 can be subject to biases including residual confounding and reverse causality.

### 4.2. Possible mechanisms

Several mechanisms may explain why obese people are at increased risk for COVID-19 infection. Inflammation and the immune system in obese individuals could play a role in relation to viral diseases. In adipose tissue there is a high production of pro-inflammatory cytokines causing chronic low-grade inflammation and immune dysregulation[35, 36]. Results from animal models revealed that the role of obesity in increasing the risk of influenza morbidity and mortality is due to the impairment of the immune response to this pathogen[37]. Green et al. proposed that hyperinsulinemia or hyperleptinemia which occurs predominantly in obese subjects may lead to a metabolic dysregulation of T cells, resulting in an impairment of the activation and function of these adaptive immune cells in response to influenza viruses[37]. In this context, Honce et al. emphasized especially the role of visceral adiposity[38]. Which metabolic and immune derangements in obese people are responsible for the increased susceptibility to COVID-19 infections should be subject of further research.

In connection with COVID-19 it is discussed that the SARS-CoV receptor ACE2 is also used by the SARS-CoV-2 spike protein (a special surface glycoprotein) as a cellular entry receptor[39]. In human tissue ACE2 is expressed in the lung, the main target site for COVID-19 infection, but also in extrapulmonary tissues including heart, kidney, and intestine. Furthermore, obesity upregulates ACE2 receptor and therefore obese subjects have larger amounts of ACE2[40]. It can be assumed that analogous to SARS-CoV[41], excessive ACE2 may competitively bind with SARS-CoV-2 not only to neutralize the virus but also to rescue cellular ACE2 activity which negatively regulates the renin-angiotensin system to protect the lung from injury[42]. Through the downregulation of ACE2 activity angiotensin II, the substrate for ACE, will accumulate and lead to increased neutrophil accumulation, increased vascular permeability, and pulmonary oedema, which will eventually cause severe lung injury. Because obesity is associated with a dysregulation of the renin-angiotensin-aldosterone system and thus among other things linked to an overexpression of angiotensin II[43], this is likely an important link between obesity and severity of COVID-19.

### 4.3. Strengths and limitations

Strengths of the study include that we performed a range of robust MR methods to conduct sensitivity analyses for different patterns of pleiotropy, investigated total and direct effects and assessed mediation mechanisms at once. The use of three body composition measures, that have different views on body fat distribution, allowed us to compare as well as differentiate between overall and abdominal fat content. Moreover, the used TFR summary level data, which based on exact measurement procedure instead of approximation, allowed us to verify the results of usually used WC-GWAS and on this way to strengthen the evidence. However, our study also has limitations. The relationship between obesity and the risk of acquiring COVID-19 disease is influenced by selection bias, because people with no, uncomplicated or milder symptoms often were not tested regarding SARS-CoV-2 infection. This causes bias towards the null hypothesis due to false negatives (type II error) and reduces therefore the power by underestimating the true causal effect. Unfortunately, neither age nor sex was available for the COVID-19 cohort, so it was not possible to perform stratified analyses. Furthermore, the present study was conducted in subjects of European ancestry and therefore the findings could not be applied to other ethnicities.

### 4.4. Conclusions

Our study is the first strengthening the evidence that overall and not abdominal obesity is causally associated with the susceptibility to and the severity of COVID-19 disease. Future research is necessary to investigate the underlying mechanisms linking obesity with COVID-19. Since the prevalence of obesity is still increasing in many countries and the probability of emerging and re-emerging infectious diseases might be high in the future[44], intensive public health interventions targeting obesity are necessary to reduce morbidity and mortality due to infectious diseases such as COVID-19.

## Data Availability

The present study is based on summary-level data that have been made publically available. Summary data from genome-wide association studies for the BMI (Pulit et al.) is available at https://zenodo.org/record/1251813#.XxgQ2J5KiUl, for WC (Shungin et al.) and BMI categories (Berndt et al.) at https://portals.broadinstitute.org/collaboration/giant/index.php/GIANT_consortium_data_files. Data for TFR (Rask-Andersen et al.) can be obtained from https://myfiles.uu.se/ssf/s/readFile/share/3993/1270878243748486898/publicLink/GWAS_summary_stats_ratios.zip[20]. The COVID-19 GWAS summary data are available at https://www.covid19hg.org/results. Summary level data for T2D can be obtained from MRC IEU OpenGWAS Project database (https://gwas.mrcieu.ac.uk/datasets/finn-a-E4_DM2/) and for CVD from http://www.cardiogramplusc4d.org/data-downloads/. In all original studies, ethical approval had been obtained.

https://myfiles.uu.se/ssf/s/readFile/share/3993/1270878243748486898/publicLink/GWAS_summary_stats_ratios.zip

https://www.covid19hg.org/results

https://gwas.mrcieu.ac.uk/datasets/finn-a-E4_DM2/

http://www.cardiogramplusc4d.org/data-downloads/

https://zenodo.org/record/1251813#.XxgQ2J5KiUl

## Author Contributions

DF analyzed the data and created the tables and figures. CM did the literature research and contributed together with DF to the data collection. Authors JL and CM reviewed and edited the paper. All authors contributed to the data interpretation.

## Data availability

The present study is based on summary-level data that have been made publically available. Summary data from genome-wide association studies for the BMI (Pulit et al.) is available at https://zenodo.org/record/1251813#.XxgQ2J5KiUl, for WC (Shungin et al.) and BMI categories (Berndt et al.) at https://portals.broadinstitute.org/collaboration/giant/index.php/GIANT_consortium_data_files. Data for TFR (Rask-Andersen et al.) can be obtained from https://myfiles.uu.se/ssf/s/readFile/share/3993/1270878243748486898/publicLink/GWAS_summary_stats_ratios.zi p[20]. The COVID-19 GWAS summary data are available at https://www.covid19hg.org/results. Summary level data for T2D can be obtained from MRC IEU OpenGWAS Project database (https://gwas.mrcieu.ac.uk/datasets/finn-a-E4_DM2/) and for CVD from http://www.cardiogramplusc4d.org/data-downloads/. In all original studies, ethical approval had been obtained.

## REFERENCES

1. Organization, W.H. Coronavirus disease (COVID-19) pandemic. 2020, October 22; Available from: https://www.who.int/emergencies/diseases/novel-coronavirus-2019.

2. Chen, J., et al., Clinical progression of patients with COVID-19 in Shanghai, China. J Infect, 2020. 80(5): p. e1–e6.

3. Aggarwal, G., et al., Association of Cardiovascular Disease With Coronavirus Disease 2019 (COVID-19) Severity: A Meta-Analysis. Curr Probl Cardiol, 2020. 45(8): p. 100617.

4. Mantovani, A., et al., Diabetes as a risk factor for greater COVID-19 severity and in-hospital death: A meta-analysis of observational studies. Nutr Metab Cardiovasc Dis, 2020. 30(8): p. 1236–1248.

5. Docherty, A.B., et al., Features of 16,749 hospitalised UK patients with COVID-19 using the ISARIC WHO Clinical Characterisation Protocol. medRxiv, 2020: p. 2020.04.23.20076042.

6. Petrilli, C.M., et al., Factors associated with hospitalization and critical illness among 4,103 patients with COVID-19 disease in New York City. medRxiv, 2020: p. 2020.04.08.20057794.

7. Williamson, E., et al., OpenSAFELY: factors associated with COVID-19-related hospital death in the linked electronic health records of 17 million adult NHS patients. medRxiv, 2020: p. 2020.05.06.20092999.

8. Du, Y., et al., Association of Body mass index (BMI) with Critical COVID-19 and in-hospital Mortality: a dose-response meta-analysis. Metabolism, 2020: p. 154373.

9. al., S.A.C.M.P.J.R.V.e., High Prevalence of Obesity in Severe Acute Respiratory Syndrome Coronavirus-2 (SARS-CoV-2) Requiring Invasive Mechanical Ventilation. Obesity (Silver Spring), 2020. 28(10): p. 1994.

10. Lighter, J., et al., Obesity in Patients Younger Than 60 Years Is a Risk Factor for COVID-19 Hospital Admission. Clin Infect Dis, 2020. 71(15): p. 896–897.

11. Yang, J., J. Hu, and C. Zhu, Obesity aggravates COVID-19: A systematic review and meta-analysis. J Med Virol, 2020.

12. Soeroto, A.Y., et al., Effect of increased BMI and obesity on the outcome of COVID-19 adult patients: A systematic review and meta-analysis. Diabetes Metab Syndr, 2020. 14(6): p. 1897–1904.

13. Mark, P.J., et al., Cardiometabolic traits, sepsis and severe covid-19 with respiratory failure: a Mendelian randomization investigation. medRxiv, 2020: p. 2020.06.18.20134676.

14. Ponsford, M.J., et al., Cardiometabolic Traits, Sepsis and Severe COVID-19: A Mendelian Randomization Investigation. Circulation, 2020.

15. Leong, A., et al., Cardiometabolic Risk Factors for COVID-19 Susceptibility and Severity: A Mendelian Randomization Analysis. medRxiv, 2020.

16. Burgess, S., C.N. Foley, and V. Zuber, Inferring Causal Relationships Between Risk Factors and Outcomes from Genome-Wide Association Study Data. Annu Rev Genomics Hum Genet, 2018. 19: p. 303–327.

17. Davies, N.M., M.V. Holmes, and G. Davey Smith, Reading Mendelian randomisation studies: a guide, glossary, and checklist for clinicians. BMJ, 2018. 362: p. k601.

18. Burgess, S. and S.G. Thompson, Mendelian randomization : methods for using genetic variants in causal estimation. Chapman & Hall/CRC interdisciplinary statistics series. 2015, Boca Raton: CRC Press, Taylor & Francis Group. xiv, 210 pages.

19. Sanderson, E., et al., An examination of multivariable Mendelian randomization in the single-sample and two-sample summary data settings. Int J Epidemiol, 2019. 48(3): p. 713–727.

20. Rask-Andersen, M., et al., Genome-wide association study of body fat distribution identifies adiposity loci and sex-specific genetic effects. Nat Commun, 2019. 10(1): p. 339.

21. Pulit, S.L., et al., Meta-analysis of genome-wide association studies for body fat distribution in 694 649 individuals of European ancestry. Hum Mol Genet, 2019. 28(1): p. 166–174.

22. Shungin, D., et al., New genetic loci link adipose and insulin biology to body fat distribution. Nature, 2015. 518(7538): p. 187–196.

23. Elsworth, B., et al., The MRC IEU OpenGWAS data infrastructure. bioRxiv, 2020: p. 2020.08.10.244293.

24. Schunkert, H., et al., Large-scale association analysis identifies 13 new susceptibility loci for coronary artery disease. Nat Genet, 2011. 43(4): p. 333–8.

25. Sanderson, E., W. Spiller, and J. Bowden, Testing and Correcting for Weak and Pleiotropic Instruments in Two-Sample Multivariable Mendelian Randomisation. bioRxiv, 2020: p. 2020.04.02.021980.

26. Berndt, S.I., et al., Genome-wide meta-analysis identifies 11 new loci for anthropometric traits and provides insights into genetic architecture. Nat Genet, 2013. 45(5): p. 501–12.

27. Initiative, C.-H.G., The COVID-19 Host Genetics Initiative, a global initiative to elucidate the role of host genetic factors in susceptibility and severity of the SARS-CoV-2 virus pandemic. Eur J Hum Genet, 2020. 28(6): p. 715–718.

28. Burgess, S., Sample size and power calculations in Mendelian randomization with a single instrumental variable and a binary outcome. Int J Epidemiol, 2014. 43(3): p. 922–9.

29. Seidu, S., et al., The impact of obesity on severe disease and mortality in people with SARS-CoV-2: A systematic review and meta-analysis. Endocrinol Diabetes Metab, 2020: p. e00176.

30. Chandarana, H., et al., Visceral adipose tissue in patients with COVID-19: risk stratification for severity. Abdom Radiol (NY), 2020.

31. Watanabe, M., et al., Visceral fat shows the strongest association with the need of intensive care in patients with COVID-19. Metabolism, 2020. 111: p. 154319.

32. Petersen, A., et al., The role of visceral adiposity in the severity of COVID-19: Highlights from a unicenter cross-sectional pilot study in Germany. Metabolism, 2020. 110: p. 154317.

33. Yang, Y., et al., Visceral Adiposity and High Intramuscular Fat Deposition Independently Predict Critical Illness in Patients with SARS-CoV-2. Obesity (Silver Spring), 2020.

34. Iacobellis, G., A.E. Malavazos, and T. Ferreira, COVID-19 Rise in Younger Adults with Obesity: Visceral Adiposity Can Predict the Risk. Obesity (Silver Spring), 2020. 28(10): p. 1795.

35. Reilly, S.M. and A.R. Saltiel, Adapting to obesity with adipose tissue inflammation. Nat Rev Endocrinol, 2017. 13(11): p. 633–643.

36. Lackey, D.E. and J.M. Olefsky, Regulation of metabolism by the innate immune system. Nat Rev Endocrinol, 2016. 12(1): p. 15–28.

37. Green, W.D. and M.A. Beck, Obesity Impairs the Adaptive Immune Response to Influenza Virus. Ann Am Thorac Soc, 2017. 14(Supplement_5): p. S406–S409.

38. Honce, R. and S. Schultz-Cherry, Impact of Obesity on Influenza A Virus Pathogenesis, Immune Response, and Evolution. Front Immunol, 2019. 10: p. 1071.

39. Hoffmann, M., H. Kleine-Weber, and S. Pohlmann, A Multibasic Cleavage Site in the Spike Protein of SARS-CoV-2 Is Essential for Infection of Human Lung Cells. Mol Cell, 2020. 78(4): p. 779–784 e5.

40. Engeli, S., et al., The adipose-tissue renin-angiotensin-aldosterone system: role in the metabolic syndrome? Int J Biochem Cell Biol, 2003. 35(6): p. 807–25.

41. Kuba, K., et al., A crucial role of angiotensin converting enzyme 2 (ACE2) in SARS coronavirus–induced lung injury. Nature Medicine, 2005. 11(8): p. 875–879.

42. Imai, Y., et al., Angiotensin-converting enzyme 2 protects from severe acute lung failure. Nature, 2005. 436(7047): p. 112–116.

43. Akoumianakis, I. and T. Filippatos, The renin-angiotensin-aldosterone system as a link between obesity and coronavirus disease 2019 severity. Obes Rev, 2020.

44. Semenza, J.C., et al., Determinants and Drivers of Infectious Disease Threat Events in Europe. Emerg Infect Dis, 2016. 22(4): p. 581–9.

