## Supplementary Material for "Impact of body composition on COVID-19 susceptibility and severity: a two-sample multivariable Mendelian randomization study"

**ABSTRACT**

**Objectives**

Recent studies suggested obesity to be a possible risk factor for COVID-19 disease in the wake of the coronavirus (SARS-CoV-2) infection. However, the causality and especially the role of body fat distribution in this context is still unclear. Thus, using a univariable as well as multivariable two-sample Mendelian randomization (MR) approach, we investigated for the first time the causal impact of body composition on the susceptibility and severity of COVID-19.

**Methods**

As indicators of overall and abdominal obesity we considered the measures body mass index (BMI), waist circumference (WC), and trunk fat ratio (TFR). Summary statistics of genome-wide association studies (GWASs) for these body composition measures were drawn from the GIANT consortium and UK Biobank, while for susceptibility and severity due to COVID-19 disease data from the COVID-19 Host Genetics Initiative was used. For the COVID-19 cohort neither age nor gender was available. Total and direct causal effect estimates were calculated using Single Nucleotide Polymorphisms (SNPs), sensitivity analyses were done applying several robust MR techniques and mediation effects of type 2 diabetes (T2D) and cardiovascular diseases (CVD) were investigated within multivariable MR analyses.

**Results**

Genetically predicted BMI was strongly associated with both, susceptibility (OR=1.31 per 1 SD increase; 95% CI: 1.15–1.50; P-value=7.3$\cdot$10^-5^) and hospitalization (OR=1.62 per 1 SD increase; 95% CI: 1.33–1.99; P-value=2.8$\cdot$10^-6^) even after adjustment for genetically predicted visceral obesity traits. These associations were neither mediated substantially by T2D nor by CVD. Finally, total but not direct effects of visceral body fat on outcomes could be detected.

**Conclusions**

This study provides strong evidence for a causal impact of overall obesity on the susceptibility and severity of COVID-19 disease. The impact of abdominal obesity was weaker and disappeared after adjustment for BMI. Therefore, obese people should be regarded as a high-risk group. Future research is necessary to investigate the underlying mechanisms linking obesity with COVID-19.

**Table of contents**

| **Single Nucleotide Polymorphisms (SNPs)** | |
| --- | --- |
| Supplementary Table S1 | Genetic instruments for body mass index |
| Supplementary Table S2 | Genetic instruments for waist circumference |
| Supplementary Table S3 | Genetic instruments for trunk fat ratio |
| **Univariable Mendelian randomization** | |
| Supplementary Table S4 | Power analysis |
| Supplementary Table S5 | Tests for detecting horizontal and directional pleiotropy |
| Supplementary Table S6 | Heterogeneity statistics |
| Supplementary Table S7 | Excluded SNPs based on Q_j_-statistics |
| **Multivariable Mendelian randomization** | |
| Supplementary Table S8 | Tests for detecting directional pleiotropy |
| Supplementary Table S9 | Heterogeneity statistics |
| **Multivariable Mendelian randomization mediation analyses** | |
| Supplementary Table S10 | Tests for detecting directional pleiotropy |
| Supplementary Table S11 | Heterogeneity statistics |
| **Sensitivity analyses** | |
| Supplementary Figure S12 | Pairwise multivariable Mendelian randomization approach |
| Supplementary Figure S13 | Mediation effects of type 2 diabetes |
| Supplementary Figure S14 | Mediation effects of cardiovascular diseases |
| Supplementary Figure S15 | Mediation effects of both, type 2 diabetes and cardiovascular diseases |

**Supplementary Table S1** Associations of genome-wide significant SNPs used as instruments for body mass index and the susceptibility as well as hospitalization due to COVID-19

|  |  |  | **COVID-19 susceptibility (n= 1079768)** | | | | **COVID-19 hospitalization (n=900687)** | | | |
| --- | --- | --- | --- | --- | --- | --- | --- | --- | --- | --- |
| **SNP** | **EA** | **OA** | **EAF** | $\boldsymbol{\beta}$ | **SE** | **P** | **EAF** | $\boldsymbol{\beta}$ | **SE** | **P** |
| rs10002111 | A | G | 0.217 | 0.014 | 0.027 | 0.604 | 0.248 | 0.068 | 0.040 | 0.087 |
| rs10033843 | A | G | 0.300 | 0.004 | 0.026 | 0.891 | 0.285 | 0.024 | 0.039 | 0.530 |
| rs10050620 | T | C | 0.361 | -0.008 | 0.024 | 0.724 | 0.369 | -0.005 | 0.035 | 0.880 |
| rs1006317 | T | G | 0.157 | -0.039 | 0.032 | 0.217 | 0.186 | -0.101 | 0.049 | 0.040 |
| rs10099330 | A | G | 0.520 | 0.001 | 0.023 | 0.960 | 0.537 | -0.038 | 0.033 | 0.252 |
| rs10101364 | T | C | 0.692 | 0.006 | 0.024 | 0.806 | 0.674 | 0.036 | 0.036 | 0.320 |
| rs10110189 | T | C | 0.177 | -0.017 | 0.036 | 0.632 | 0.186 | -0.015 | 0.052 | 0.780 |
| rs10132280 | A | C | 0.334 | 0.016 | 0.024 | 0.490 | 0.332 | 0.047 | 0.035 | 0.181 |
| rs10145749 | T | C | 0.209 | 0.018 | 0.031 | 0.559 | 0.217 | 0.123 | 0.047 | 0.009 |
| rs10168563 | A | G | 0.670 | -0.030 | 0.025 | 0.216 | 0.641 | -0.003 | 0.037 | 0.935 |
| rs10169594 | T | C | 0.651 | 0.026 | 0.023 | 0.268 | 0.630 | 0.024 | 0.034 | 0.481 |
| rs10182181 | A | G | 0.505 | 0.010 | 0.022 | 0.662 | 0.524 | 0.012 | 0.033 | 0.715 |
| rs10185199 | A | G | 0.322 | -0.013 | 0.026 | 0.625 | 0.341 | 0.035 | 0.038 | 0.357 |
| rs10261050 | T | C | 0.452 | 0.035 | 0.022 | 0.121 | 0.450 | 0.021 | 0.033 | 0.531 |
| rs1048637 | T | G | 0.544 | 0.020 | 0.025 | 0.436 | 0.528 | 0.006 | 0.034 | 0.857 |
| rs10499694 | A | G | 0.527 | -0.022 | 0.022 | 0.321 | 0.522 | -0.012 | 0.033 | 0.714 |
| rs10506971 | A | G | 0.512 | -0.003 | 0.022 | 0.893 | 0.523 | -0.016 | 0.033 | 0.635 |
| rs10518694 | A | C | 0.161 | 0.007 | 0.032 | 0.838 | 0.181 | -0.010 | 0.048 | 0.840 |
| rs10733051 | A | G | 0.534 | -0.018 | 0.022 | 0.414 | 0.516 | -0.062 | 0.034 | 0.067 |
| rs10733682 | A | G | 0.483 | 0.001 | 0.026 | 0.974 | 0.509 | 0.009 | 0.034 | 0.790 |
| rs10741329 | A | G | 0.647 | 0.037 | 0.024 | 0.119 | 0.643 | 0.035 | 0.035 | 0.320 |
| rs10742752 | T | C | 0.371 | -0.006 | 0.023 | 0.797 | 0.387 | -0.044 | 0.034 | 0.195 |
| rs1075901 | T | C | 0.443 | -0.015 | 0.022 | 0.513 | 0.456 | -0.030 | 0.033 | 0.368 |
| rs10761785 | T | G | 0.510 | 0.028 | 0.022 | 0.203 | 0.519 | 0.025 | 0.033 | 0.451 |
| rs10772983 | T | C | 0.510 | 0.011 | 0.022 | 0.616 | 0.514 | 0.032 | 0.033 | 0.328 |
| rs10779751 | A | G | 0.302 | 0.039 | 0.024 | 0.106 | 0.300 | 0.062 | 0.037 | 0.092 |
| rs10811868 | A | G | 0.331 | -0.013 | 0.024 | 0.584 | 0.348 | -0.006 | 0.035 | 0.869 |
| rs10829164 | T | C | 0.202 | 0.029 | 0.031 | 0.350 | 0.230 | 0.065 | 0.046 | 0.158 |
| rs10842240 | C | G | 0.181 | 0.075 | 0.033 | 0.022 | 0.196 | 0.058 | 0.048 | 0.229 |
| rs10858334 | C | G | 0.850 | -0.001 | 0.034 | 0.984 | 0.825 | -0.060 | 0.048 | 0.216 |
| rs10864728 | A | G | 0.449 | 0.010 | 0.022 | 0.661 | 0.439 | -0.021 | 0.033 | 0.523 |
| rs10909880 | T | C | 0.416 | -0.001 | 0.022 | 0.976 | 0.432 | 0.055 | 0.033 | 0.096 |
| rs10920336 | A | G | 0.550 | -0.024 | 0.022 | 0.273 | 0.548 | 0.007 | 0.033 | 0.842 |
| rs10920678 | A | G | 0.426 | -0.037 | 0.022 | 0.094 | 0.436 | -0.040 | 0.033 | 0.229 |
| rs10929925 | A | C | 0.490 | -0.037 | 0.023 | 0.099 | 0.472 | -0.048 | 0.033 | 0.151 |
| rs10938397 | A | G | 0.592 | 0.007 | 0.022 | 0.749 | 0.570 | 0.015 | 0.033 | 0.661 |
| rs10942267 | A | G | 0.679 | 0.018 | 0.024 | 0.461 | 0.689 | 0.025 | 0.036 | 0.476 |
| rs10961649 | T | C | 0.322 | 0.018 | 0.024 | 0.454 | 0.339 | -0.004 | 0.035 | 0.897 |
| rs10962550 | C | G | 0.205 | 0.025 | 0.029 | 0.393 | 0.224 | 0.007 | 0.042 | 0.875 |
| rs10968114 | A | C | 0.547 | -0.013 | 0.022 | 0.564 | 0.535 | 0.034 | 0.034 | 0.318 |
| rs10975933 | C | G | 0.655 | 0.021 | 0.024 | 0.388 | 0.633 | 0.030 | 0.035 | 0.398 |
| rs10992867 | A | G | 0.322 | 0.021 | 0.025 | 0.405 | 0.344 | 0.023 | 0.037 | 0.532 |
| rs11001259 | A | T | 0.169 | 0.014 | 0.031 | 0.660 | 0.218 | 0.033 | 0.046 | 0.483 |
| rs11030618 | T | C | 0.584 | 0.018 | 0.022 | 0.428 | 0.565 | 0.017 | 0.033 | 0.618 |
| rs11044430 | A | T | 0.741 | 0.040 | 0.034 | 0.238 | 0.733 | 0.070 | 0.046 | 0.125 |
| rs11046972 | T | C | 0.105 | 0.037 | 0.041 | 0.360 | 0.140 | 0.050 | 0.058 | 0.390 |
| rs11060853 | A | G | 0.599 | 0.044 | 0.023 | 0.051 | 0.579 | 0.018 | 0.034 | 0.587 |
| rs11066188 | A | G | 0.328 | 0.006 | 0.023 | 0.795 | 0.367 | 0.005 | 0.034 | 0.893 |
| rs11078883 | C | G | 0.627 | -0.025 | 0.023 | 0.284 | 0.626 | 0.018 | 0.035 | 0.610 |
| rs11105839 | A | T | 0.401 | -0.036 | 0.023 | 0.116 | 0.396 | -0.071 | 0.034 | 0.038 |
| rs11115176 | T | C | 0.772 | 0.014 | 0.026 | 0.607 | 0.742 | -0.009 | 0.039 | 0.810 |
| rs11121210 | T | C | 0.408 | -0.029 | 0.022 | 0.192 | 0.419 | -0.072 | 0.034 | 0.033 |
| rs11128021 | A | G | 0.160 | 0.053 | 0.031 | 0.088 | 0.184 | 0.069 | 0.047 | 0.142 |
| rs11150911 | A | C | 0.357 | 0.016 | 0.025 | 0.533 | 0.353 | -0.022 | 0.038 | 0.557 |
| rs11165643 | T | C | 0.527 | 0.013 | 0.022 | 0.546 | 0.552 | 0.012 | 0.033 | 0.716 |
| rs11170468 | A | C | 0.777 | -0.005 | 0.027 | 0.865 | 0.760 | -0.005 | 0.039 | 0.905 |
| rs11218510 | A | G | 0.354 | 0.021 | 0.023 | 0.358 | 0.377 | 0.033 | 0.034 | 0.336 |
| rs11246136 | A | C | 0.119 | -0.060 | 0.039 | 0.127 | 0.153 | -0.094 | 0.058 | 0.103 |
| rs112646560 | T | C | 0.219 | 0.003 | 0.027 | 0.906 | 0.254 | 0.087 | 0.042 | 0.036 |
| rs1126930 | C | G | 0.058 | 0.080 | 0.066 | 0.225 | 0.094 | 0.121 | 0.105 | 0.245 |
| rs11525873 | T | C | 0.876 | 0.073 | 0.039 | 0.060 | 0.850 | 0.062 | 0.057 | 0.280 |
| rs11538 | A | G | 0.845 | -0.019 | 0.031 | 0.533 | 0.814 | 0.029 | 0.047 | 0.531 |
| rs11577094 | T | C | 0.088 | 0.051 | 0.042 | 0.230 | 0.118 | 0.096 | 0.068 | 0.159 |
| rs11594179 | T | C | 0.215 | -0.017 | 0.027 | 0.537 | 0.232 | -0.001 | 0.039 | 0.988 |
| rs1159692 | A | C | 0.495 | -0.040 | 0.023 | 0.085 | 0.496 | -0.051 | 0.034 | 0.134 |
| rs11614340 | T | C | 0.746 | 0.013 | 0.026 | 0.600 | 0.715 | -0.023 | 0.037 | 0.530 |
| rs11615578 | T | C | 0.232 | -0.005 | 0.027 | 0.841 | 0.252 | -0.011 | 0.040 | 0.786 |
| rs11633626 | A | C | 0.627 | -0.022 | 0.023 | 0.336 | 0.620 | 0.017 | 0.035 | 0.628 |
| rs11636611 | T | C | 0.478 | 0.046 | 0.022 | 0.040 | 0.490 | 0.053 | 0.033 | 0.108 |
| rs116374395 | A | G | 0.065 | 0.042 | 0.056 | 0.449 | 0.097 | 0.019 | 0.085 | 0.821 |
| rs11649864 | A | G | 0.108 | 0.025 | 0.040 | 0.536 | 0.134 | -0.011 | 0.059 | 0.853 |
| rs11655587 | T | C | 0.340 | 0.005 | 0.023 | 0.822 | 0.348 | 0.031 | 0.035 | 0.370 |
| rs11672660 | T | C | 0.207 | -0.008 | 0.028 | 0.778 | 0.233 | -0.070 | 0.042 | 0.093 |
| rs11692326 | T | C | 0.220 | -0.029 | 0.027 | 0.285 | 0.246 | 0.009 | 0.041 | 0.829 |
| rs11695013 | T | C | 0.596 | -0.044 | 0.023 | 0.057 | 0.618 | -0.062 | 0.034 | 0.071 |
| rs11713193 | A | G | 0.450 | 0.016 | 0.022 | 0.462 | 0.437 | 0.042 | 0.033 | 0.201 |
| rs11739877 | T | C | 0.622 | -0.008 | 0.023 | 0.720 | 0.603 | -0.056 | 0.034 | 0.102 |
| rs11757278 | T | C | 0.705 | 0.050 | 0.025 | 0.042 | 0.682 | 0.069 | 0.037 | 0.059 |
| rs11772246 | T | C | 0.803 | 0.031 | 0.029 | 0.280 | 0.767 | 0.039 | 0.042 | 0.355 |
| rs11773362 | T | C | 0.326 | 0.016 | 0.024 | 0.500 | 0.355 | -0.014 | 0.035 | 0.701 |
| rs11782074 | T | G | 0.327 | 0.010 | 0.024 | 0.665 | 0.357 | 0.009 | 0.035 | 0.807 |
| rs118081010 | T | C | 0.056 | -0.045 | 0.079 | 0.570 | 0.083 | -0.201 | 0.117 | 0.086 |
| rs11882409 | A | C | 0.277 | 0.029 | 0.025 | 0.253 | 0.287 | 0.034 | 0.037 | 0.357 |
| rs11902450 | T | C | 0.142 | 0.024 | 0.037 | 0.504 | 0.152 | 0.026 | 0.054 | 0.622 |
| rs11915371 | A | C | 0.774 | -0.042 | 0.027 | 0.120 | 0.763 | -0.066 | 0.041 | 0.106 |
| rs11919665 | A | T | 0.373 | -0.036 | 0.028 | 0.192 | 0.388 | -0.012 | 0.036 | 0.748 |
| rs11921432 | T | C | 0.881 | 0.015 | 0.037 | 0.677 | 0.851 | -0.001 | 0.054 | 0.981 |
| rs12072739 | A | G | 0.752 | -0.037 | 0.025 | 0.141 | 0.732 | -0.011 | 0.040 | 0.782 |
| rs12098284 | T | C | 0.157 | -0.004 | 0.033 | 0.896 | 0.171 | -0.048 | 0.049 | 0.329 |
| rs12140153 | T | G | 0.103 | -0.051 | 0.040 | 0.205 | 0.133 | 0.002 | 0.066 | 0.981 |
| rs12150665 | T | C | 0.625 | 0.001 | 0.023 | 0.973 | 0.592 | 0.015 | 0.033 | 0.645 |
| rs12259464 | A | G | 0.428 | 0.006 | 0.023 | 0.777 | 0.443 | 0.027 | 0.033 | 0.419 |
| rs12282785 | A | C | 0.259 | -0.006 | 0.027 | 0.829 | 0.264 | 0.028 | 0.039 | 0.480 |
| rs12286929 | A | G | 0.490 | -0.049 | 0.025 | 0.045 | 0.501 | -0.065 | 0.033 | 0.053 |
| rs12334877 | A | G | 0.235 | 0.019 | 0.028 | 0.496 | 0.251 | 0.053 | 0.041 | 0.196 |
| rs12364470 | T | G | 0.829 | 0.025 | 0.032 | 0.435 | 0.795 | 0.007 | 0.048 | 0.889 |
| rs12369179 | T | C | 0.085 | -0.008 | 0.046 | 0.870 | 0.122 | 0.034 | 0.069 | 0.623 |
| rs12386885 | T | C | 0.144 | 0.009 | 0.036 | 0.802 | 0.178 | -0.015 | 0.048 | 0.756 |
| rs12421848 | A | G | 0.429 | 0.014 | 0.025 | 0.587 | 0.434 | 0.023 | 0.034 | 0.512 |
| rs12429545 | A | G | 0.158 | 0.046 | 0.033 | 0.162 | 0.182 | 0.025 | 0.049 | 0.611 |
| rs12439632 | C | G | 0.195 | 0.017 | 0.031 | 0.579 | 0.218 | 0.034 | 0.049 | 0.478 |
| rs12448257 | A | G | 0.228 | -0.022 | 0.027 | 0.416 | 0.239 | 0.009 | 0.040 | 0.824 |
| rs12462975 | A | G | 0.298 | -0.017 | 0.024 | 0.490 | 0.321 | -0.009 | 0.036 | 0.798 |
| rs12527426 | A | G | 0.364 | 0.037 | 0.025 | 0.140 | 0.361 | 0.041 | 0.036 | 0.264 |
| rs12591120 | T | C | 0.732 | -0.014 | 0.026 | 0.587 | 0.725 | 0.006 | 0.038 | 0.874 |
| rs12602912 | T | C | 0.218 | -0.005 | 0.028 | 0.862 | 0.249 | -0.009 | 0.041 | 0.820 |
| rs12611148 | A | C | 0.167 | -0.012 | 0.031 | 0.701 | 0.189 | 0.019 | 0.046 | 0.680 |
| rs12628051 | T | C | 0.646 | 0.001 | 0.024 | 0.953 | 0.623 | -0.022 | 0.035 | 0.519 |
| rs12628891 | T | C | 0.287 | 0.021 | 0.025 | 0.402 | 0.306 | 0.068 | 0.038 | 0.069 |
| rs12636480 | T | G | 0.338 | 0.011 | 0.023 | 0.626 | 0.354 | 0.049 | 0.034 | 0.157 |
| rs12652212 | A | G | 0.576 | 0.014 | 0.022 | 0.534 | 0.577 | -0.023 | 0.033 | 0.478 |
| rs1268065 | A | G | 0.534 | 0.012 | 0.023 | 0.585 | 0.508 | -0.037 | 0.033 | 0.262 |
| rs12680842 | A | G | 0.668 | 0.024 | 0.024 | 0.312 | 0.639 | 0.040 | 0.035 | 0.253 |
| rs12681792^a^ | A | C | 0.225 | -0.019 | 0.028 | 0.494 |  |  |  |  |
| rs12692596 | T | C | 0.363 | 0.009 | 0.023 | 0.687 | 0.378 | -0.009 | 0.034 | 0.799 |
| rs12714199 | T | C | 0.593 | 0.002 | 0.023 | 0.918 | 0.579 | 0.007 | 0.033 | 0.834 |
| rs12765914 | T | C | 0.107 | -0.023 | 0.039 | 0.566 | 0.142 | -0.029 | 0.058 | 0.612 |
| rs12868881 | A | T | 0.390 | -0.010 | 0.023 | 0.657 | 0.416 | -0.020 | 0.034 | 0.549 |
| rs12888545 | A | G | 0.780 | -0.013 | 0.026 | 0.614 | 0.753 | -0.036 | 0.039 | 0.359 |
| rs12912198 | T | C | 0.262 | -0.062 | 0.026 | 0.016 | 0.279 | -0.087 | 0.038 | 0.022 |
| rs12914623 | C | G | 0.284 | 0.011 | 0.025 | 0.675 | 0.286 | -0.023 | 0.038 | 0.546 |
| rs12922346 | C | G | 0.267 | 0.014 | 0.026 | 0.594 | 0.275 | -0.008 | 0.039 | 0.840 |
| rs12926250 | T | G | 0.107 | 0.059 | 0.038 | 0.122 | 0.134 | -0.015 | 0.055 | 0.781 |
| rs1293037 | T | C | 0.743 | -0.012 | 0.027 | 0.664 | 0.728 | -0.004 | 0.039 | 0.913 |
| rs12939549 | A | G | 0.585 | -0.015 | 0.022 | 0.512 | 0.586 | 0.013 | 0.033 | 0.697 |
| rs1296328 | A | C | 0.518 | -0.039 | 0.023 | 0.092 | 0.503 | -0.026 | 0.033 | 0.443 |
| rs12981256 | A | G | 0.477 | 0.015 | 0.023 | 0.506 | 0.499 | 0.005 | 0.035 | 0.883 |
| rs12987009 | A | T | 0.605 | -0.012 | 0.023 | 0.610 | 0.588 | -0.002 | 0.034 | 0.960 |
| rs13021737 | A | G | 0.186 | -0.027 | 0.030 | 0.370 | 0.220 | 0.014 | 0.043 | 0.752 |
| rs13033310 | A | G | 0.264 | -0.016 | 0.026 | 0.530 | 0.279 | -0.004 | 0.038 | 0.907 |
| rs1304549 | A | G | 0.269 | -0.053 | 0.027 | 0.053 | 0.259 | -0.022 | 0.040 | 0.580 |
| rs13107325 | T | C | 0.079 | 0.068 | 0.049 | 0.164 | 0.119 | 0.061 | 0.062 | 0.328 |
| rs13110266 | A | G | 0.410 | -0.033 | 0.023 | 0.141 | 0.422 | -0.043 | 0.034 | 0.201 |
| rs13174863 | A | G | 0.844 | -0.049 | 0.031 | 0.119 | 0.810 | -0.031 | 0.045 | 0.490 |
| rs13186194 | T | C | 0.615 | 0.019 | 0.023 | 0.416 | 0.613 | -0.006 | 0.035 | 0.856 |
| rs13191362 | A | G | 0.868 | 0.019 | 0.034 | 0.581 | 0.844 | 0.079 | 0.050 | 0.114 |
| rs13240600 | A | G | 0.729 | -0.031 | 0.031 | 0.322 | 0.739 | -0.060 | 0.045 | 0.179 |
| rs13245051 | A | G | 0.516 | 0.030 | 0.023 | 0.183 | 0.509 | 0.056 | 0.034 | 0.096 |
| rs13263601 | A | C | 0.689 | -0.047 | 0.024 | 0.047 | 0.679 | 0.007 | 0.035 | 0.849 |
| rs1327259 | A | G | 0.583 | 0.027 | 0.023 | 0.234 | 0.565 | 0.006 | 0.034 | 0.855 |
| rs13296413 | T | C | 0.371 | -0.014 | 0.023 | 0.531 | 0.377 | 0.000 | 0.034 | 0.995 |
| rs13298487 | T | C | 0.606 | -0.020 | 0.024 | 0.392 | 0.616 | -0.037 | 0.036 | 0.304 |
| rs1346841 | A | G | 0.444 | 0.018 | 0.023 | 0.423 | 0.431 | -0.032 | 0.034 | 0.351 |
| rs1350430 | T | C | 0.528 | -0.017 | 0.022 | 0.437 | 0.539 | -0.045 | 0.033 | 0.176 |
| rs1356506 | T | C | 0.650 | -0.022 | 0.023 | 0.352 | 0.627 | 0.003 | 0.035 | 0.931 |
| rs1358980 | T | C | 0.478 | -0.010 | 0.023 | 0.669 | 0.498 | -0.011 | 0.033 | 0.734 |
| rs1383592 | A | G | 0.254 | -0.013 | 0.026 | 0.611 | 0.273 | -0.035 | 0.039 | 0.371 |
| rs1409818 | T | C | 0.149 | -0.006 | 0.035 | 0.861 | 0.166 | -0.057 | 0.052 | 0.268 |
| rs1412235 | C | G | 0.282 | -0.001 | 0.025 | 0.954 | 0.304 | 0.034 | 0.037 | 0.361 |
| rs1421334 | A | C | 0.504 | -0.013 | 0.023 | 0.576 | 0.484 | -0.037 | 0.033 | 0.274 |
| rs1437842 | A | G | 0.499 | 0.030 | 0.022 | 0.174 | 0.474 | 0.010 | 0.033 | 0.765 |
| rs1441264 | A | G | 0.631 | 0.005 | 0.023 | 0.819 | 0.612 | -0.029 | 0.034 | 0.399 |
| rs1451077 | A | G | 0.539 | -0.047 | 0.023 | 0.038 | 0.542 | -0.068 | 0.034 | 0.043 |
| rs147568678 | T | C | 0.781 | 0.005 | 0.028 | 0.868 | 0.743 | -0.020 | 0.041 | 0.631 |
| rs1477199 | A | G | 0.830 | -0.041 | 0.032 | 0.194 | 0.813 | -0.046 | 0.046 | 0.324 |
| rs1492014 | T | C | 0.570 | -0.001 | 0.023 | 0.963 | 0.586 | -0.040 | 0.034 | 0.239 |
| rs1492767 | T | C | 0.432 | 0.020 | 0.022 | 0.381 | 0.453 | 0.033 | 0.033 | 0.321 |
| rs1501673 | A | G | 0.170 | 0.021 | 0.033 | 0.522 | 0.190 | 0.015 | 0.047 | 0.754 |
| rs150215901 | A | T | 0.068 | 0.093 | 0.057 | 0.103 | 0.119 | 0.012 | 0.082 | 0.882 |
| rs1522569 | T | G | 0.803 | -0.010 | 0.029 | 0.723 | 0.791 | 0.044 | 0.042 | 0.295 |
| rs1559673 | A | C | 0.932 | -0.036 | 0.070 | 0.610 | 0.895 | -0.146 | 0.109 | 0.179 |
| rs156201 | C | G | 0.683 | 0.006 | 0.029 | 0.831 | 0.681 | 0.008 | 0.039 | 0.834 |
| rs16851483 | T | G | 0.097 | -0.054 | 0.044 | 0.224 | 0.125 | 0.000 | 0.066 | 1.000 |
| rs16906838 | T | C | 0.091 | -0.023 | 0.048 | 0.634 | 0.114 | -0.057 | 0.071 | 0.427 |
| rs1700082 | C | G | 0.591 | 0.047 | 0.024 | 0.053 | 0.610 | 0.026 | 0.035 | 0.462 |
| rs17020497 | A | G | 0.158 | -0.005 | 0.033 | 0.887 | 0.168 | -0.023 | 0.047 | 0.624 |
| rs17024393 | T | C | 0.943 | -0.120 | 0.065 | 0.066 | 0.914 | -0.174 | 0.108 | 0.109 |
| rs17066856 | T | C | 0.836 | 0.028 | 0.038 | 0.456 | 0.828 | 0.045 | 0.055 | 0.418 |
| rs17094222 | T | C | 0.797 | -0.014 | 0.028 | 0.621 | 0.762 | -0.036 | 0.040 | 0.370 |
| rs17182027 | A | G | 0.498 | 0.041 | 0.022 | 0.068 | 0.506 | 0.059 | 0.034 | 0.078 |
| rs17207196 | T | C | 0.388 | -0.005 | 0.023 | 0.821 | 0.411 | -0.009 | 0.035 | 0.790 |
| rs1721447 | T | G | 0.503 | -0.034 | 0.022 | 0.132 | 0.507 | -0.034 | 0.033 | 0.313 |
| rs17367750 | T | C | 0.322 | 0.028 | 0.024 | 0.241 | 0.339 | 0.075 | 0.036 | 0.034 |
| rs17405603 | A | T | 0.736 | -0.001 | 0.025 | 0.963 | 0.711 | 0.007 | 0.038 | 0.861 |
| rs17405819 | T | C | 0.709 | -0.016 | 0.024 | 0.518 | 0.682 | -0.059 | 0.036 | 0.102 |
| rs1750307 | A | T | 0.385 | -0.003 | 0.025 | 0.915 | 0.394 | -0.010 | 0.035 | 0.764 |
| rs17544384 | T | C | 0.801 | 0.001 | 0.027 | 0.968 | 0.774 | -0.014 | 0.040 | 0.722 |
| rs17636031 | T | C | 0.728 | -0.018 | 0.026 | 0.501 | 0.698 | -0.033 | 0.037 | 0.384 |
| rs17681451 | A | G | 0.102 | 0.070 | 0.041 | 0.084 | 0.131 | 0.104 | 0.061 | 0.086 |
| rs17724992 | A | G | 0.715 | -0.021 | 0.025 | 0.401 | 0.693 | -0.006 | 0.037 | 0.870 |
| rs17783165 | T | C | 0.623 | -0.020 | 0.024 | 0.407 | 0.616 | -0.028 | 0.035 | 0.422 |
| rs17806224 | A | G | 0.180 | -0.021 | 0.029 | 0.466 | 0.213 | 0.013 | 0.043 | 0.756 |
| rs17814208 | A | G | 0.755 | -0.007 | 0.026 | 0.787 | 0.731 | -0.024 | 0.039 | 0.529 |
| rs1799923 | A | G | 0.170 | -0.037 | 0.035 | 0.282 | 0.177 | -0.053 | 0.053 | 0.315 |
| rs1808629 | A | G | 0.612 | 0.011 | 0.024 | 0.652 | 0.631 | 0.027 | 0.035 | 0.441 |
| rs185350 | T | C | 0.471 | -0.001 | 0.022 | 0.966 | 0.486 | 0.022 | 0.033 | 0.501 |
| rs1860561 | A | G | 0.210 | -0.002 | 0.028 | 0.947 | 0.237 | -0.025 | 0.041 | 0.546 |
| rs1877875 | T | C | 0.413 | -0.018 | 0.022 | 0.428 | 0.410 | -0.019 | 0.033 | 0.576 |
| rs1884389 | T | C | 0.429 | -0.033 | 0.022 | 0.147 | 0.433 | -0.046 | 0.033 | 0.168 |
| rs1884897 | A | G | 0.390 | -0.019 | 0.023 | 0.414 | 0.382 | -0.022 | 0.034 | 0.517 |
| rs1927790 | T | C | 0.562 | -0.019 | 0.022 | 0.405 | 0.560 | -0.021 | 0.033 | 0.532 |
| rs1928295 | T | C | 0.557 | -0.020 | 0.022 | 0.369 | 0.549 | -0.034 | 0.033 | 0.306 |
| rs1941213 | A | C | 0.706 | 0.023 | 0.025 | 0.369 | 0.690 | 0.051 | 0.038 | 0.175 |
| rs1941696 | A | G | 0.511 | 0.010 | 0.022 | 0.645 | 0.518 | -0.009 | 0.033 | 0.782 |
| rs1945160 | A | G | 0.354 | -0.031 | 0.023 | 0.183 | 0.367 | -0.047 | 0.034 | 0.168 |
| rs1948080 | T | G | 0.648 | -0.044 | 0.023 | 0.060 | 0.630 | -0.053 | 0.035 | 0.126 |
| rs194809 | A | G | 0.212 | -0.028 | 0.028 | 0.323 | 0.234 | -0.045 | 0.042 | 0.277 |
| rs1951455 | T | C | 0.289 | -0.002 | 0.025 | 0.941 | 0.305 | 0.000 | 0.036 | 0.993 |
| rs1958898 | C | G | 0.204 | -0.063 | 0.029 | 0.028 | 0.239 | -0.040 | 0.043 | 0.350 |
| rs1965529 | A | G | 0.752 | -0.033 | 0.027 | 0.219 | 0.732 | -0.045 | 0.040 | 0.263 |
| rs197374 | T | C | 0.413 | -0.012 | 0.022 | 0.597 | 0.405 | -0.024 | 0.034 | 0.478 |
| rs1999433 | T | C | 0.442 | 0.002 | 0.023 | 0.944 | 0.446 | 0.038 | 0.034 | 0.258 |
| rs2007518 | A | G | 0.605 | -0.016 | 0.022 | 0.471 | 0.600 | -0.024 | 0.033 | 0.473 |
| rs2047648 | A | T | 0.755 | 0.020 | 0.026 | 0.438 | 0.725 | -0.018 | 0.037 | 0.632 |
| rs2051559 | T | C | 0.845 | -0.016 | 0.033 | 0.618 | 0.824 | -0.047 | 0.051 | 0.352 |
| rs2053682 | A | C | 0.688 | 0.007 | 0.024 | 0.781 | 0.665 | 0.015 | 0.035 | 0.671 |
| rs2058527 | T | G | 0.269 | -0.032 | 0.025 | 0.202 | 0.292 | -0.038 | 0.038 | 0.306 |
| rs2064044 | A | C | 0.777 | 0.005 | 0.027 | 0.843 | 0.747 | -0.032 | 0.042 | 0.446 |
| rs2065418 | T | G | 0.661 | 0.051 | 0.023 | 0.028 | 0.642 | 0.020 | 0.034 | 0.550 |
| rs2066295 | A | G | 0.771 | 0.033 | 0.026 | 0.213 | 0.752 | 0.045 | 0.039 | 0.253 |
| rs2074314 | T | C | 0.656 | -0.013 | 0.023 | 0.577 | 0.614 | -0.025 | 0.034 | 0.465 |
| rs2108719 | A | G | 0.706 | -0.022 | 0.026 | 0.397 | 0.697 | 0.014 | 0.038 | 0.713 |
| rs2112347 | T | G | 0.588 | -0.040 | 0.023 | 0.079 | 0.589 | -0.062 | 0.034 | 0.068 |
| rs2119753 | A | G | 0.639 | -0.006 | 0.023 | 0.796 | 0.622 | -0.013 | 0.035 | 0.697 |
| rs2120710 | A | G | 0.655 | -0.024 | 0.024 | 0.314 | 0.628 | -0.002 | 0.035 | 0.962 |
| rs2134858 | T | C | 0.506 | 0.021 | 0.022 | 0.341 | 0.504 | -0.029 | 0.033 | 0.369 |
| rs213518 | T | C | 0.840 | -0.044 | 0.032 | 0.175 | 0.825 | 0.000 | 0.048 | 0.994 |
| rs214249 | T | G | 0.616 | 0.009 | 0.023 | 0.695 | 0.615 | 0.025 | 0.034 | 0.466 |
| rs215669 | A | G | 0.541 | -0.026 | 0.023 | 0.266 | 0.568 | -0.016 | 0.034 | 0.640 |
| rs217433 | T | C | 0.793 | 0.004 | 0.028 | 0.900 | 0.774 | -0.006 | 0.043 | 0.883 |
| rs217669 | T | C | 0.697 | 0.020 | 0.026 | 0.435 | 0.706 | -0.007 | 0.038 | 0.843 |
| rs2192158 | A | G | 0.480 | -0.011 | 0.022 | 0.621 | 0.482 | 0.036 | 0.033 | 0.278 |
| rs2196618 | A | G | 0.306 | 0.025 | 0.026 | 0.338 | 0.303 | 0.007 | 0.038 | 0.847 |
| rs2206277 | T | C | 0.202 | 0.016 | 0.029 | 0.578 | 0.229 | 0.011 | 0.042 | 0.789 |
| rs2228213 | A | G | 0.317 | -0.025 | 0.024 | 0.281 | 0.350 | -0.032 | 0.034 | 0.352 |
| rs2228552 | T | G | 0.594 | -0.051 | 0.022 | 0.022 | 0.608 | -0.070 | 0.034 | 0.041 |
| rs2238799 | A | G | 0.589 | 0.051 | 0.023 | 0.027 | 0.567 | 0.076 | 0.034 | 0.025 |
| rs2241423 | A | G | 0.284 | 0.020 | 0.026 | 0.444 | 0.278 | -0.017 | 0.038 | 0.657 |
| rs2246012 | T | C | 0.812 | 0.020 | 0.030 | 0.505 | 0.783 | -0.001 | 0.046 | 0.981 |
| rs2257791 | A | G | 0.720 | 0.027 | 0.026 | 0.291 | 0.715 | 0.064 | 0.038 | 0.093 |
| rs225882 | T | C | 0.693 | -0.007 | 0.025 | 0.783 | 0.672 | 0.027 | 0.037 | 0.462 |
| rs2267958 | A | G | 0.469 | -0.019 | 0.023 | 0.408 | 0.488 | -0.031 | 0.035 | 0.374 |
| rs2271046 | A | T | 0.649 | -0.012 | 0.025 | 0.619 | 0.665 | -0.025 | 0.037 | 0.506 |
| rs2271189 | A | G | 0.348 | -0.029 | 0.023 | 0.206 | 0.369 | -0.028 | 0.034 | 0.421 |
| rs2273175 | T | C | 0.703 | -0.023 | 0.025 | 0.359 | 0.673 | -0.089 | 0.036 | 0.014 |
| rs2275003 | A | G | 0.498 | 0.012 | 0.022 | 0.585 | 0.500 | -0.017 | 0.033 | 0.603 |
| rs2281819 | A | T | 0.244 | 0.009 | 0.026 | 0.723 | 0.268 | 0.064 | 0.039 | 0.105 |
| rs2283006 | A | G | 0.540 | -0.005 | 0.023 | 0.840 | 0.508 | -0.031 | 0.034 | 0.362 |
| rs2283093 | T | C | 0.212 | -0.006 | 0.028 | 0.821 | 0.236 | -0.004 | 0.041 | 0.929 |
| rs2289379 | T | C | 0.368 | -0.015 | 0.023 | 0.508 | 0.398 | -0.008 | 0.035 | 0.817 |
| rs2342892 | T | G | 0.444 | -0.012 | 0.023 | 0.601 | 0.464 | -0.002 | 0.033 | 0.962 |
| rs2357760 | A | G | 0.658 | -0.015 | 0.024 | 0.532 | 0.632 | -0.054 | 0.035 | 0.117 |
| rs2365389 | T | C | 0.485 | -0.031 | 0.022 | 0.174 | 0.472 | -0.039 | 0.033 | 0.249 |
| rs2396625 | A | T | 0.382 | 0.020 | 0.023 | 0.382 | 0.399 | -0.013 | 0.034 | 0.706 |
| rs2400414 | T | C | 0.367 | -0.008 | 0.022 | 0.719 | 0.365 | -0.009 | 0.034 | 0.782 |
| rs2423668 | T | C | 0.466 | 0.011 | 0.023 | 0.636 | 0.458 | 0.015 | 0.034 | 0.658 |
| rs2436728 | A | G | 0.378 | -0.022 | 0.023 | 0.343 | 0.407 | 0.019 | 0.033 | 0.568 |
| rs2439823 | A | G | 0.462 | 0.016 | 0.022 | 0.481 | 0.490 | -0.001 | 0.033 | 0.978 |
| rs2466103 | T | G | 0.718 | 0.034 | 0.024 | 0.164 | 0.702 | 0.008 | 0.036 | 0.815 |
| rs2470893 | T | C | 0.245 | 0.010 | 0.025 | 0.684 | 0.268 | 0.072 | 0.038 | 0.057 |
| rs2503185 | A | G | 0.474 | -0.013 | 0.022 | 0.546 | 0.484 | 0.005 | 0.033 | 0.878 |
| rs2513999 | A | G | 0.205 | 0.022 | 0.031 | 0.487 | 0.201 | 0.026 | 0.046 | 0.575 |
| rs2600226 | T | C | 0.617 | 0.000 | 0.024 | 0.988 | 0.609 | 0.009 | 0.035 | 0.790 |
| rs2605603 | A | G | 0.513 | -0.036 | 0.022 | 0.102 | 0.516 | -0.069 | 0.033 | 0.035 |
| rs2612576 | A | T | 0.334 | -0.054 | 0.025 | 0.032 | 0.321 | -0.093 | 0.037 | 0.012 |
| rs2622274 | T | G | 0.453 | -0.017 | 0.023 | 0.458 | 0.438 | -0.019 | 0.034 | 0.574 |
| rs264941 | A | C | 0.467 | 0.036 | 0.022 | 0.108 | 0.472 | 0.015 | 0.033 | 0.641 |
| rs2707183 | T | G | 0.528 | -0.018 | 0.023 | 0.426 | 0.532 | -0.007 | 0.033 | 0.838 |
| rs2712665 | T | C | 0.726 | -0.010 | 0.024 | 0.691 | 0.698 | -0.027 | 0.036 | 0.446 |
| rs2715423 | A | G | 0.250 | -0.049 | 0.025 | 0.054 | 0.274 | -0.034 | 0.038 | 0.371 |
| rs273512 | T | C | 0.410 | -0.014 | 0.023 | 0.524 | 0.416 | -0.002 | 0.034 | 0.945 |
| rs2744974 | T | C | 0.369 | 0.009 | 0.024 | 0.714 | 0.356 | -0.035 | 0.035 | 0.316 |
| rs274628 | A | C | 0.325 | -0.015 | 0.023 | 0.514 | 0.345 | 0.001 | 0.035 | 0.970 |
| rs2777768 | A | G | 0.747 | 0.036 | 0.026 | 0.162 | 0.735 | 0.056 | 0.037 | 0.137 |
| rs2820295 | A | G | 0.317 | 0.000 | 0.023 | 0.996 | 0.327 | -0.014 | 0.035 | 0.691 |
| rs2832283 | A | G | 0.217 | 0.012 | 0.027 | 0.655 | 0.246 | 0.029 | 0.041 | 0.476 |
| rs28350 | A | G | 0.193 | 0.026 | 0.029 | 0.371 | 0.228 | 0.071 | 0.042 | 0.091 |
| rs28489620 | A | G | 0.263 | 0.035 | 0.025 | 0.159 | 0.285 | 0.034 | 0.038 | 0.362 |
| rs2861089 | A | T | 0.380 | 0.003 | 0.023 | 0.899 | 0.392 | 0.058 | 0.034 | 0.086 |
| rs2861685 | T | C | 0.585 | -0.034 | 0.023 | 0.128 | 0.578 | -0.069 | 0.033 | 0.038 |
| rs2862996 | T | G | 0.672 | 0.023 | 0.024 | 0.351 | 0.657 | 0.015 | 0.035 | 0.672 |
| rs2875762 | C | G | 0.280 | -0.006 | 0.026 | 0.820 | 0.278 | 0.069 | 0.038 | 0.069 |
| rs2907948 | A | G | 0.231 | -0.010 | 0.026 | 0.700 | 0.257 | -0.015 | 0.038 | 0.692 |
| rs2910026 | T | C | 0.719 | 0.010 | 0.025 | 0.705 | 0.717 | -0.007 | 0.037 | 0.847 |
| rs2962334 | T | G | 0.054 | 0.135 | 0.079 | 0.086 | 0.079 | 0.055 | 0.115 | 0.632 |
| rs2984618 | T | G | 0.445 | -0.007 | 0.022 | 0.745 | 0.438 | 0.007 | 0.034 | 0.841 |
| rs3019466 | T | C | 0.170 | 0.000 | 0.031 | 0.992 | 0.188 | -0.026 | 0.046 | 0.564 |
| rs305256 | T | C | 0.234 | 0.043 | 0.028 | 0.118 | 0.273 | -0.032 | 0.042 | 0.447 |
| rs3101336 | T | C | 0.387 | 0.010 | 0.025 | 0.696 | 0.375 | 0.024 | 0.035 | 0.487 |
| rs3115667 | T | C | 0.257 | 0.007 | 0.029 | 0.805 | 0.255 | -0.033 | 0.040 | 0.408 |
| rs312750 | A | G | 0.536 | -0.030 | 0.023 | 0.180 | 0.505 | -0.050 | 0.033 | 0.132 |
| rs321237 | A | G | 0.741 | 0.014 | 0.025 | 0.576 | 0.718 | -0.016 | 0.037 | 0.677 |
| rs326893 | T | C | 0.546 | 0.015 | 0.023 | 0.518 | 0.566 | 0.055 | 0.034 | 0.109 |
| rs329651 | T | G | 0.825 | -0.040 | 0.028 | 0.156 | 0.793 | 0.020 | 0.041 | 0.621 |
| rs337637 | A | G | 0.323 | -0.021 | 0.024 | 0.372 | 0.349 | -0.051 | 0.035 | 0.147 |
| rs339991 | A | G | 0.435 | -0.038 | 0.023 | 0.091 | 0.444 | -0.019 | 0.033 | 0.570 |
| rs34184235 | T | C | 0.388 | -0.003 | 0.023 | 0.888 | 0.420 | -0.004 | 0.033 | 0.910 |
| rs34234296 | A | G | 0.364 | -0.039 | 0.023 | 0.092 | 0.373 | -0.076 | 0.034 | 0.026 |
| rs34517439 | A | C | 0.108 | 0.044 | 0.036 | 0.226 | 0.134 | 0.084 | 0.058 | 0.146 |
| rs34811474 | A | G | 0.211 | -0.034 | 0.028 | 0.222 | 0.253 | -0.059 | 0.042 | 0.168 |
| rs349088 | A | C | 0.502 | -0.003 | 0.022 | 0.886 | 0.521 | -0.034 | 0.033 | 0.297 |
| rs35408866 | A | G | 0.144 | 0.034 | 0.034 | 0.320 | 0.172 | 0.052 | 0.048 | 0.282 |
| rs35483388 | T | C | 0.389 | -0.019 | 0.024 | 0.429 | 0.393 | -0.034 | 0.035 | 0.322 |
| rs355777 | C | G | 0.410 | 0.018 | 0.023 | 0.428 |  |  |  |  |
| rs35867081 | A | G | 0.485 | 0.000 | 0.022 | 0.988 | 0.481 | -0.008 | 0.033 | 0.805 |
| rs35949039 | T | G | 0.126 | -0.055 | 0.037 | 0.136 | 0.157 | 0.008 | 0.056 | 0.885 |
| rs3764625 | T | G | 0.400 | 0.003 | 0.023 | 0.878 | 0.426 | -0.050 | 0.034 | 0.134 |
| rs3764835 | A | G | 0.155 | 0.005 | 0.031 | 0.871 | 0.185 | 0.096 | 0.046 | 0.039 |
| rs3770890 | T | G | 0.942 | 0.028 | 0.087 | 0.749 | 0.862 | -0.097 | 0.133 | 0.464 |
| rs3796432 | T | G | 0.353 | -0.018 | 0.023 | 0.444 | 0.366 | -0.001 | 0.034 | 0.967 |
| rs3806114 | A | G | 0.714 | -0.008 | 0.025 | 0.743 | 0.689 | -0.040 | 0.037 | 0.277 |
| rs3808477 | T | C | 0.253 | -0.024 | 0.026 | 0.359 | 0.280 | -0.016 | 0.039 | 0.677 |
| rs3814883 | T | C | 0.430 | 0.009 | 0.023 | 0.700 | 0.440 | -0.043 | 0.034 | 0.199 |
| rs3825061 | T | C | 0.373 | -0.021 | 0.023 | 0.350 | 0.375 | -0.014 | 0.034 | 0.688 |
| rs3902951 | T | G | 0.739 | -0.015 | 0.028 | 0.592 | 0.721 | -0.005 | 0.041 | 0.904 |
| rs3914628 | T | C | 0.827 | -0.009 | 0.031 | 0.784 | 0.805 | 0.024 | 0.046 | 0.604 |
| rs3923783 | A | C | 0.183 | 0.018 | 0.029 | 0.543 | 0.207 | -0.003 | 0.045 | 0.952 |
| rs39654 | A | G | 0.437 | 0.043 | 0.023 | 0.057 | 0.444 | 0.038 | 0.034 | 0.256 |
| rs40067 | A | G | 0.218 | -0.023 | 0.029 | 0.441 | 0.243 | -0.027 | 0.043 | 0.532 |
| rs4017425 | T | C | 0.480 | -0.002 | 0.022 | 0.942 | 0.462 | -0.003 | 0.033 | 0.927 |
| rs4097319 | T | G | 0.591 | 0.016 | 0.023 | 0.493 | 0.566 | 0.012 | 0.034 | 0.715 |
| rs4148155 | A | G | 0.883 | 0.000 | 0.037 | 0.996 | 0.857 | 0.001 | 0.056 | 0.990 |
| rs4240673 | T | C | 0.468 | 0.041 | 0.023 | 0.078 | 0.487 | 0.036 | 0.033 | 0.275 |
| rs4256980 | C | G | 0.400 | 0.006 | 0.023 | 0.795 | 0.404 | 0.020 | 0.034 | 0.553 |
| rs427943 | A | C | 0.438 | 0.013 | 0.023 | 0.562 | 0.452 | -0.030 | 0.033 | 0.371 |
| rs4286488 | A | G | 0.763 | 0.012 | 0.027 | 0.656 | 0.736 | -0.001 | 0.039 | 0.988 |
| rs4303732 | T | C | 0.653 | 0.048 | 0.023 | 0.039 | 0.634 | 0.052 | 0.034 | 0.131 |
| rs4307239 | A | G | 0.508 | -0.001 | 0.023 | 0.959 | 0.518 | -0.025 | 0.033 | 0.452 |
| rs4390583 | A | C | 0.591 | 0.004 | 0.023 | 0.868 | 0.588 | 0.030 | 0.034 | 0.372 |
| rs4430672 | T | C | 0.225 | -0.015 | 0.028 | 0.595 | 0.237 | -0.011 | 0.041 | 0.795 |
| rs4482463 | A | C | 0.860 | -0.049 | 0.040 | 0.224 | 0.843 | -0.126 | 0.059 | 0.033 |
| rs4517716 | C | G | 0.724 | 0.021 | 0.027 | 0.434 | 0.722 | -0.018 | 0.040 | 0.649 |
| rs45486197 | A | G | 0.081 | -0.055 | 0.049 | 0.260 | 0.118 | -0.012 | 0.074 | 0.873 |
| rs459552 | A | T | 0.769 | 0.045 | 0.027 | 0.096 | 0.736 | 0.025 | 0.040 | 0.526 |
| rs4653017 | T | C | 0.670 | 0.018 | 0.023 | 0.434 | 0.648 | 0.038 | 0.035 | 0.282 |
| rs4655141^a^ | T | C | 0.757 | 0.032 | 0.028 | 0.257 |  |  |  |  |
| rs4671328 | T | G | 0.443 | -0.043 | 0.032 | 0.182 | 0.442 | -0.021 | 0.048 | 0.667 |
| rs4700608 | T | C | 0.511 | 0.030 | 0.023 | 0.189 | 0.489 | 0.030 | 0.033 | 0.357 |
| rs4721089 | T | C | 0.742 | 0.009 | 0.026 | 0.734 | 0.712 | 0.027 | 0.038 | 0.490 |
| rs4740383 | A | G | 0.454 | -0.011 | 0.026 | 0.676 | 0.435 | -0.009 | 0.035 | 0.797 |
| rs4740619 | T | C | 0.525 | 0.015 | 0.022 | 0.504 | 0.509 | 0.032 | 0.033 | 0.324 |
| rs474605 | A | G | 0.424 | 0.024 | 0.023 | 0.296 | 0.454 | 0.044 | 0.033 | 0.178 |
| rs4771218 | A | G | 0.573 | 0.002 | 0.023 | 0.921 | 0.591 | -0.011 | 0.033 | 0.750 |
| rs478707 | T | C | 0.275 | 0.059 | 0.027 | 0.030 | 0.281 | 0.046 | 0.040 | 0.246 |
| rs4812405 | A | C | 0.083 | 0.031 | 0.051 | 0.539 | 0.123 | 0.035 | 0.075 | 0.642 |
| rs4813619 | T | G | 0.555 | -0.031 | 0.023 | 0.171 | 0.529 | -0.036 | 0.034 | 0.296 |
| rs4858193 | T | C | 0.759 | 0.022 | 0.026 | 0.401 | 0.733 | 0.011 | 0.039 | 0.770 |
| rs4864201 | T | C | 0.433 | 0.013 | 0.024 | 0.578 | 0.411 | -0.001 | 0.035 | 0.987 |
| rs4865796 | A | G | 0.692 | -0.038 | 0.024 | 0.114 | 0.671 | -0.056 | 0.036 | 0.121 |
| rs4880341 | T | C | 0.553 | 0.003 | 0.022 | 0.904 | 0.548 | -0.026 | 0.033 | 0.427 |
| rs4900714 | T | G | 0.548 | -0.015 | 0.024 | 0.532 | 0.530 | 0.021 | 0.034 | 0.541 |
| rs4906263 | C | G | 0.657 | -0.023 | 0.024 | 0.340 | 0.637 | -0.028 | 0.035 | 0.419 |
| rs4921301 | T | C | 0.238 | -0.032 | 0.027 | 0.247 | 0.264 | -0.002 | 0.040 | 0.955 |
| rs4970991 | T | C | 0.263 | 0.008 | 0.026 | 0.753 | 0.270 | -0.014 | 0.041 | 0.722 |
| rs4973618 | A | G | 0.652 | -0.024 | 0.023 | 0.299 | 0.637 | 0.010 | 0.035 | 0.779 |
| rs4981693 | A | G | 0.735 | -0.007 | 0.026 | 0.800 | 0.719 | 0.026 | 0.040 | 0.515 |
| rs4986044 | T | C | 0.514 | -0.002 | 0.022 | 0.914 | 0.498 | -0.055 | 0.034 | 0.100 |
| rs4988235^a^ | A | G | 0.487 | -0.048 | 0.028 | 0.085 |  |  |  |  |
| rs543874 | A | G | 0.794 | -0.004 | 0.028 | 0.893 | 0.782 | -0.063 | 0.043 | 0.146 |
| rs56133507 | T | G | 0.800 | -0.013 | 0.033 | 0.691 | 0.777 | -0.043 | 0.043 | 0.324 |
| rs56151256 | A | C | 0.765 | 0.048 | 0.027 | 0.072 | 0.749 | 0.050 | 0.039 | 0.208 |
| rs56161855 | A | T | 0.839 | -0.043 | 0.038 | 0.256 | 0.800 | -0.032 | 0.049 | 0.518 |
| rs56211164 | A | G | 0.250 | 0.010 | 0.026 | 0.697 | 0.270 | -0.009 | 0.039 | 0.814 |
| rs562664 | T | C | 0.180 | -0.008 | 0.033 | 0.799 | 0.216 | -0.007 | 0.044 | 0.866 |
| rs570463 | A | C | 0.330 | 0.035 | 0.024 | 0.142 | 0.336 | -0.023 | 0.035 | 0.525 |
| rs587271 | T | C | 0.702 | -0.004 | 0.024 | 0.875 | 0.665 | -0.030 | 0.037 | 0.416 |
| rs592483 | T | C | 0.595 | -0.006 | 0.023 | 0.794 | 0.566 | -0.001 | 0.034 | 0.988 |
| rs59302296 | A | T | 0.108 | 0.006 | 0.039 | 0.881 | 0.135 | -0.044 | 0.057 | 0.435 |
| rs6010784 | T | C | 0.430 | 0.007 | 0.023 | 0.744 | 0.475 | 0.002 | 0.033 | 0.958 |
| rs61740466 | A | G | 0.249 | 0.015 | 0.025 | 0.544 | 0.274 | 0.005 | 0.038 | 0.900 |
| rs61813324 | T | C | 0.136 | 0.027 | 0.034 | 0.439 | 0.180 | 0.065 | 0.052 | 0.214 |
| rs61828641 | A | G | 0.134 | 0.033 | 0.035 | 0.352 | 0.162 | 0.122 | 0.053 | 0.022 |
| rs61983990 | A | G | 0.098 | -0.041 | 0.046 | 0.368 | 0.139 | 0.085 | 0.066 | 0.196 |
| rs62176243 | A | T | 0.759 | 0.066 | 0.026 | 0.012 | 0.733 | 0.055 | 0.039 | 0.161 |
| rs6265 | T | C | 0.192 | -0.027 | 0.029 | 0.353 | 0.223 | -0.053 | 0.041 | 0.199 |
| rs6443750 | T | C | 0.180 | -0.028 | 0.030 | 0.360 | 0.232 | -0.030 | 0.046 | 0.514 |
| rs6445258 | T | C | 0.218 | -0.026 | 0.028 | 0.366 | 0.248 | 0.001 | 0.043 | 0.988 |
| rs6463489 | T | C | 0.120 | -0.021 | 0.037 | 0.563 | 0.150 | 0.039 | 0.057 | 0.488 |
| rs6470144 | T | G | 0.627 | -0.004 | 0.023 | 0.865 | 0.627 | -0.006 | 0.035 | 0.857 |
| rs6493498 | T | C | 0.460 | 0.031 | 0.025 | 0.210 | 0.469 | 0.043 | 0.034 | 0.199 |
| rs6496248 | A | T | 0.645 | -0.026 | 0.026 | 0.308 | 0.631 | -0.054 | 0.035 | 0.125 |
| rs6500208 | A | G | 0.243 | -0.022 | 0.028 | 0.422 | 0.253 | -0.008 | 0.042 | 0.839 |
| rs650198 | T | C | 0.707 | -0.022 | 0.025 | 0.382 | 0.678 | 0.009 | 0.036 | 0.813 |
| rs6512302 | C | G | 0.720 | -0.020 | 0.026 | 0.437 | 0.707 | 0.020 | 0.039 | 0.618 |
| rs6539064 | C | G | 0.691 | -0.016 | 0.025 | 0.535 | 0.679 | -0.031 | 0.037 | 0.396 |
| rs6545714 | A | G | 0.622 | -0.020 | 0.023 | 0.373 | 0.615 | -0.035 | 0.034 | 0.301 |
| rs6556301 | T | G | 0.355 | -0.008 | 0.024 | 0.725 | 0.375 | 0.012 | 0.035 | 0.734 |
| rs6567160 | T | C | 0.735 | -0.015 | 0.029 | 0.593 | 0.725 | 0.004 | 0.039 | 0.924 |
| rs657452 | A | G | 0.432 | 0.035 | 0.022 | 0.108 | 0.431 | 0.038 | 0.034 | 0.255 |
| rs6591407 | A | C | 0.183 | 0.066 | 0.029 | 0.026 | 0.209 | 0.031 | 0.043 | 0.466 |
| rs6607337 | T | C | 0.328 | 0.031 | 0.024 | 0.199 | 0.322 | 0.031 | 0.036 | 0.383 |
| rs6656785 | A | G | 0.607 | -0.048 | 0.022 | 0.030 | 0.589 | -0.050 | 0.034 | 0.136 |
| rs66595146 | A | C | 0.612 | 0.020 | 0.023 | 0.390 | 0.595 | 0.080 | 0.034 | 0.020 |
| rs6661316 | T | C | 0.562 | -0.002 | 0.022 | 0.939 | 0.549 | -0.040 | 0.033 | 0.224 |
| rs6696828 | C | G | 0.327 | 0.001 | 0.024 | 0.980 | 0.309 | -0.022 | 0.037 | 0.549 |
| rs6710091 | C | G | 0.678 | 0.010 | 0.024 | 0.664 | 0.661 | -0.048 | 0.035 | 0.174 |
| rs6716898 | A | G | 0.521 | 0.021 | 0.023 | 0.351 | 0.490 | 0.042 | 0.034 | 0.211 |
| rs6720868 | T | C | 0.353 | -0.013 | 0.024 | 0.584 | 0.361 | -0.024 | 0.035 | 0.497 |
| rs6725931 | T | C | 0.796 | -0.010 | 0.030 | 0.735 | 0.793 | -0.020 | 0.043 | 0.641 |
| rs6783054 | A | C | 0.522 | 0.009 | 0.022 | 0.701 | 0.523 | -0.042 | 0.033 | 0.204 |
| rs6803161 | T | C | 0.434 | 0.011 | 0.023 | 0.615 | 0.434 | 0.077 | 0.033 | 0.021 |
| rs6804842 | A | G | 0.463 | -0.011 | 0.022 | 0.626 | 0.463 | 0.008 | 0.033 | 0.804 |
| rs6808814 | T | C | 0.737 | 0.004 | 0.026 | 0.890 | 0.717 | -0.008 | 0.039 | 0.837 |
| rs6850421 | A | G | 0.491 | 0.056 | 0.023 | 0.014 | 0.499 | 0.072 | 0.033 | 0.032 |
| rs6864049 | A | G | 0.427 | 0.030 | 0.023 | 0.189 | 0.442 | 0.017 | 0.033 | 0.603 |
| rs687339 | T | C | 0.752 | 0.019 | 0.026 | 0.467 | 0.740 | 0.014 | 0.039 | 0.727 |
| rs6882366 | T | C | 0.403 | 0.019 | 0.023 | 0.398 | 0.400 | 0.040 | 0.034 | 0.243 |
| rs6886072 | T | C | 0.438 | -0.008 | 0.022 | 0.725 | 0.458 | -0.041 | 0.033 | 0.210 |
| rs6888194 | T | C | 0.804 | -0.005 | 0.031 | 0.877 | 0.808 | 0.018 | 0.047 | 0.694 |
| rs6890310 | A | G | 0.288 | -0.015 | 0.025 | 0.557 | 0.296 | -0.041 | 0.037 | 0.258 |
| rs6893539 | A | C | 0.686 | -0.029 | 0.024 | 0.240 | 0.677 | -0.040 | 0.037 | 0.281 |
| rs6909685 | T | C | 0.377 | 0.010 | 0.023 | 0.660 | 0.390 | 0.002 | 0.035 | 0.951 |
| rs6915002 | T | C | 0.446 | -0.004 | 0.023 | 0.861 | 0.434 | -0.028 | 0.034 | 0.410 |
| rs6921533 | T | C | 0.358 | 0.001 | 0.024 | 0.952 | 0.351 | 0.015 | 0.036 | 0.683 |
| rs6922607 | A | G | 0.783 | 0.007 | 0.028 | 0.793 | 0.764 | 0.069 | 0.041 | 0.089 |
| rs6950388 | A | G | 0.672 | 0.036 | 0.028 | 0.187 | 0.701 | 0.061 | 0.041 | 0.134 |
| rs6973656 | A | G | 0.605 | -0.020 | 0.023 | 0.389 | 0.602 | 0.017 | 0.033 | 0.605 |
| rs698147 | A | G | 0.441 | -0.034 | 0.023 | 0.139 | 0.466 | -0.060 | 0.033 | 0.071 |
| rs7024334 | T | G | 0.229 | -0.037 | 0.027 | 0.161 | 0.257 | -0.076 | 0.039 | 0.049 |
| rs7070670 | T | C | 0.306 | -0.014 | 0.024 | 0.579 | 0.321 | -0.025 | 0.036 | 0.489 |
| rs7084454 | A | G | 0.342 | -0.014 | 0.024 | 0.555 | 0.343 | 0.006 | 0.035 | 0.852 |
| rs7102454 | T | C | 0.701 | -0.032 | 0.024 | 0.181 | 0.679 | -0.039 | 0.035 | 0.270 |
| rs7124681 | A | C | 0.389 | 0.033 | 0.023 | 0.141 | 0.405 | 0.010 | 0.033 | 0.768 |
| rs7138803 | A | G | 0.364 | 0.015 | 0.023 | 0.512 | 0.376 | 0.028 | 0.034 | 0.414 |
| rs7144011 | T | G | 0.212 | 0.003 | 0.028 | 0.915 | 0.230 | 0.015 | 0.041 | 0.711 |
| rs7161194 | A | G | 0.369 | -0.021 | 0.025 | 0.396 | 0.371 | -0.026 | 0.036 | 0.465 |
| rs7171864 | A | G | 0.668 | -0.038 | 0.024 | 0.116 | 0.658 | -0.035 | 0.036 | 0.320 |
| rs7172627 | A | G | 0.495 | 0.023 | 0.022 | 0.297 | 0.509 | 0.035 | 0.033 | 0.293 |
| rs7206608 | C | G | 0.674 | -0.048 | 0.024 | 0.046 | 0.665 | -0.029 | 0.036 | 0.415 |
| rs7245985 | T | G | 0.770 | -0.047 | 0.027 | 0.088 | 0.754 | -0.044 | 0.039 | 0.265 |
| rs72649373 | T | C | 0.863 | 0.038 | 0.033 | 0.242 | 0.830 | 0.061 | 0.049 | 0.210 |
| rs72673947 | A | G | 0.859 | -0.022 | 0.040 | 0.582 | 0.873 | 0.021 | 0.070 | 0.763 |
| rs72757415 | T | G | 0.210 | 0.037 | 0.028 | 0.180 | 0.221 | -0.018 | 0.043 | 0.676 |
| rs73213484 | A | T | 0.815 | -0.012 | 0.031 | 0.706 | 0.788 | -0.004 | 0.045 | 0.921 |
| rs73225274 | A | G | 0.830 | -0.020 | 0.033 | 0.542 | 0.808 | -0.049 | 0.048 | 0.302 |
| rs7323 | C | G | 0.250 | -0.014 | 0.025 | 0.590 | 0.273 | -0.006 | 0.038 | 0.879 |
| rs7357754 | A | G | 0.495 | -0.025 | 0.022 | 0.265 | 0.511 | 0.005 | 0.033 | 0.877 |
| rs73985439 | A | C | 0.668 | -0.033 | 0.024 | 0.173 | 0.656 | -0.045 | 0.036 | 0.203 |
| rs742748 | T | C | 0.564 | -0.007 | 0.023 | 0.748 | 0.576 | -0.032 | 0.033 | 0.334 |
| rs74887628 | A | G | 0.067 | 0.133 | 0.075 | 0.076 | 0.143 | 0.098 | 0.119 | 0.410 |
| rs7498665 | A | G | 0.652 | 0.003 | 0.023 | 0.913 | 0.636 | -0.013 | 0.035 | 0.700 |
| rs750090 | T | C | 0.605 | -0.042 | 0.023 | 0.072 | 0.608 | -0.009 | 0.035 | 0.803 |
| rs7512146 | T | G | 0.485 | 0.007 | 0.022 | 0.748 | 0.504 | 0.002 | 0.033 | 0.954 |
| rs7534091 | A | G | 0.735 | -0.010 | 0.025 | 0.697 | 0.711 | 0.016 | 0.038 | 0.675 |
| rs7561278 | T | C | 0.765 | -0.022 | 0.027 | 0.430 | 0.740 | 0.000 | 0.041 | 0.993 |
| rs756717 | A | G | 0.366 | 0.002 | 0.023 | 0.937 | 0.385 | -0.015 | 0.035 | 0.668 |
| rs7588437 | A | G | 0.343 | -0.010 | 0.023 | 0.669 | 0.356 | 0.021 | 0.034 | 0.538 |
| rs7593917 | A | G | 0.474 | -0.002 | 0.022 | 0.946 | 0.466 | -0.079 | 0.033 | 0.018 |
| rs7599312 | A | G | 0.277 | -0.020 | 0.025 | 0.422 | 0.282 | -0.035 | 0.038 | 0.363 |
| rs7616009 | A | G | 0.170 | -0.016 | 0.032 | 0.615 | 0.196 | -0.076 | 0.046 | 0.101 |
| rs7631156 | A | G | 0.303 | 0.011 | 0.024 | 0.633 | 0.322 | -0.008 | 0.035 | 0.826 |
| rs7640424 | T | C | 0.275 | -0.013 | 0.025 | 0.601 | 0.301 | -0.016 | 0.036 | 0.656 |
| rs765125 | T | C | 0.526 | -0.037 | 0.023 | 0.101 | 0.536 | -0.038 | 0.033 | 0.246 |
| rs765875 | T | C | 0.473 | -0.009 | 0.022 | 0.678 | 0.491 | 0.005 | 0.033 | 0.870 |
| rs76638898 | A | G | 0.058 | 0.041 | 0.077 | 0.598 | 0.106 | 0.101 | 0.114 | 0.378 |
| rs7678054 | A | G | 0.466 | 0.014 | 0.022 | 0.532 | 0.465 | 0.007 | 0.033 | 0.831 |
| rs768023 | A | G | 0.546 | 0.017 | 0.023 | 0.453 | 0.553 | 0.020 | 0.034 | 0.551 |
| rs76942203 | A | G | 0.100 | -0.019 | 0.044 | 0.666 | 0.124 | -0.022 | 0.067 | 0.739 |
| rs7696649 | A | G | 0.272 | 0.014 | 0.025 | 0.577 | 0.293 | -0.003 | 0.037 | 0.944 |
| rs7713317 | A | G | 0.703 | 0.015 | 0.024 | 0.533 | 0.679 | 0.043 | 0.036 | 0.235 |
| rs7715256 | T | G | 0.566 | 0.003 | 0.022 | 0.892 | 0.554 | -0.044 | 0.033 | 0.185 |
| rs77165542 | T | C | 0.059 | 0.023 | 0.065 | 0.722 | 0.109 | -0.114 | 0.092 | 0.216 |
| rs7727781 | T | C | 0.482 | 0.021 | 0.022 | 0.345 | 0.502 | 0.030 | 0.033 | 0.366 |
| rs7730004 | T | C | 0.651 | 0.024 | 0.024 | 0.306 | 0.643 | 0.043 | 0.036 | 0.227 |
| rs7734385 | A | G | 0.392 | 0.028 | 0.023 | 0.227 | 0.419 | 0.019 | 0.034 | 0.576 |
| rs77432547 | A | G | 0.735 | -0.005 | 0.028 | 0.859 | 0.707 | 0.034 | 0.039 | 0.382 |
| rs7760482 | A | G | 0.612 | 0.022 | 0.023 | 0.340 | 0.598 | 0.034 | 0.034 | 0.318 |
| rs7774 | A | C | 0.397 | 0.010 | 0.027 | 0.717 | 0.400 | 0.009 | 0.046 | 0.851 |
| rs7802342 | T | G | 0.689 | 0.027 | 0.024 | 0.271 | 0.684 | 0.054 | 0.036 | 0.134 |
| rs7842934 | T | C | 0.861 | 0.012 | 0.041 | 0.768 | 0.854 | 0.012 | 0.063 | 0.845 |
| rs7861160 | T | C | 0.551 | 0.050 | 0.023 | 0.028 | 0.553 | 0.073 | 0.034 | 0.029 |
| rs7893571 | T | G | 0.696 | 0.036 | 0.025 | 0.150 | 0.668 | 0.024 | 0.035 | 0.493 |
| rs7899106 | A | G | 0.920 | -0.080 | 0.049 | 0.104 | 0.895 | -0.053 | 0.076 | 0.487 |
| rs7903146 | T | C | 0.312 | 0.023 | 0.024 | 0.332 | 0.325 | 0.043 | 0.035 | 0.226 |
| rs7907470 | A | G | 0.854 | -0.045 | 0.038 | 0.235 | 0.834 | 0.000 | 0.060 | 0.997 |
| rs79113395 | A | G | 0.273 | 0.018 | 0.036 | 0.613 | 0.262 | 0.041 | 0.054 | 0.446 |
| rs79186842 | A | G | 0.874 | 0.030 | 0.035 | 0.384 | 0.845 | 0.003 | 0.053 | 0.958 |
| rs7944782 | T | G | 0.482 | -0.026 | 0.023 | 0.250 | 0.488 | -0.050 | 0.034 | 0.136 |
| rs7975187 | A | G | 0.744 | -0.023 | 0.027 | 0.397 | 0.736 | -0.063 | 0.039 | 0.110 |
| rs79780963 | T | C | 0.191 | -0.014 | 0.039 | 0.728 | 0.155 | -0.045 | 0.056 | 0.428 |
| rs79906980 | T | C | 0.156 | -0.046 | 0.031 | 0.144 | 0.178 | -0.013 | 0.047 | 0.774 |
| rs805412 | A | G | 0.461 | 0.013 | 0.023 | 0.565 | 0.457 | 0.014 | 0.033 | 0.666 |
| rs8057911 | T | C | 0.257 | 0.005 | 0.027 | 0.847 | 0.269 | 0.036 | 0.040 | 0.364 |
| rs8065172 | A | G | 0.264 | 0.027 | 0.026 | 0.287 | 0.285 | 0.071 | 0.038 | 0.062 |
| rs8097672 | A | T | 0.817 | -0.037 | 0.032 | 0.246 | 0.806 | -0.124 | 0.047 | 0.008 |
| rs8122855 | A | G | 0.317 | 0.042 | 0.024 | 0.076 | 0.338 | 0.039 | 0.035 | 0.269 |
| rs8126575 | T | G | 0.825 | 0.049 | 0.033 | 0.137 | 0.796 | 0.028 | 0.048 | 0.553 |
| rs8134638 | T | C | 0.618 | 0.004 | 0.023 | 0.855 | 0.591 | -0.014 | 0.035 | 0.693 |
| rs8181823 | A | C | 0.260 | -0.034 | 0.026 | 0.202 | 0.276 | -0.082 | 0.039 | 0.036 |
| rs845084 | A | G | 0.292 | -0.002 | 0.025 | 0.929 | 0.311 | 0.007 | 0.037 | 0.849 |
| rs852056 | T | C | 0.317 | -0.038 | 0.026 | 0.139 | 0.323 | -0.011 | 0.038 | 0.771 |
| rs865809 | A | G | 0.244 | -0.001 | 0.027 | 0.976 | 0.277 | 0.032 | 0.040 | 0.428 |
| rs879620 | T | C | 0.528 | 0.011 | 0.023 | 0.632 | 0.549 | -0.009 | 0.034 | 0.780 |
| rs889398 | T | C | 0.388 | -0.017 | 0.023 | 0.458 | 0.402 | -0.012 | 0.034 | 0.726 |
| rs891387 | T | C | 0.500 | 0.033 | 0.022 | 0.136 | 0.502 | 0.030 | 0.033 | 0.362 |
| rs895330 | C | G | 0.745 | 0.012 | 0.028 | 0.679 | 0.759 | 0.085 | 0.042 | 0.040 |
| rs900144 | T | C | 0.490 | -0.019 | 0.023 | 0.394 | 0.511 | -0.041 | 0.033 | 0.223 |
| rs9168 | A | C | 0.280 | -0.022 | 0.028 | 0.424 | 0.292 | -0.031 | 0.038 | 0.417 |
| rs925018 | C | G | 0.637 | -0.011 | 0.024 | 0.646 | 0.650 | -0.050 | 0.035 | 0.155 |
| rs9294260 | A | G | 0.500 | 0.020 | 0.022 | 0.382 | 0.472 | 0.060 | 0.033 | 0.071 |
| rs9299 | T | C | 0.586 | -0.006 | 0.023 | 0.781 | 0.604 | 0.027 | 0.034 | 0.430 |
| rs930295 | A | C | 0.172 | -0.012 | 0.030 | 0.690 | 0.199 | -0.003 | 0.045 | 0.951 |
| rs9304665 | A | T | 0.688 | -0.027 | 0.026 | 0.307 | 0.673 | -0.030 | 0.040 | 0.450 |
| rs9320823 | T | C | 0.359 | 0.005 | 0.023 | 0.827 | 0.379 | -0.014 | 0.034 | 0.683 |
| rs935166 | A | G | 0.446 | 0.005 | 0.023 | 0.836 | 0.463 | -0.009 | 0.034 | 0.784 |
| rs9370410^a^ | A | G | 0.724 | 0.053 | 0.025 | 0.030 |  |  |  |  |
| rs9375702 | T | C | 0.632 | -0.001 | 0.025 | 0.978 | 0.641 | -0.007 | 0.036 | 0.841 |
| rs942066 | A | G | 0.397 | 0.006 | 0.023 | 0.792 | 0.407 | 0.024 | 0.034 | 0.476 |
| rs9458814 | T | C | 0.737 | 0.022 | 0.026 | 0.402 | 0.720 | 0.011 | 0.039 | 0.778 |
| rs946824 | T | C | 0.173 | -0.049 | 0.032 | 0.128 | 0.196 | -0.078 | 0.048 | 0.107 |
| rs9478496 | T | C | 0.834 | 0.018 | 0.031 | 0.553 | 0.805 | -0.036 | 0.045 | 0.421 |
| rs9512648 | A | G | 0.456 | -0.009 | 0.023 | 0.704 | 0.466 | -0.019 | 0.033 | 0.565 |
| rs9522183 | T | G | 0.502 | -0.006 | 0.022 | 0.791 | 0.507 | -0.017 | 0.033 | 0.614 |
| rs9527895 | T | C | 0.830 | -0.027 | 0.030 | 0.362 | 0.801 | -0.072 | 0.044 | 0.098 |
| rs9531786 | C | G | 0.404 | -0.014 | 0.023 | 0.542 | 0.403 | -0.018 | 0.034 | 0.597 |
| rs9569777 | T | G | 0.210 | -0.007 | 0.029 | 0.802 | 0.234 | 0.015 | 0.045 | 0.734 |
| rs9595908 | T | C | 0.645 | -0.037 | 0.023 | 0.107 | 0.625 | -0.073 | 0.034 | 0.033 |
| rs9599161 | T | C | 0.569 | 0.032 | 0.023 | 0.164 | 0.582 | 0.042 | 0.034 | 0.217 |
| rs9603697 | T | C | 0.331 | 0.021 | 0.024 | 0.368 | 0.363 | -0.016 | 0.035 | 0.642 |
| rs962796 | T | C | 0.242 | 0.020 | 0.027 | 0.453 | 0.246 | 0.056 | 0.040 | 0.165 |
| rs9816226 | A | T | 0.207 | -0.030 | 0.028 | 0.295 | 0.225 | 0.005 | 0.042 | 0.907 |
| rs9818122 | T | C | 0.782 | -0.034 | 0.027 | 0.213 | 0.770 | -0.031 | 0.040 | 0.437 |
| rs9826775 | A | G | 0.827 | -0.032 | 0.031 | 0.304 | 0.807 | -0.046 | 0.046 | 0.322 |
| rs9827823 | T | C | 0.799 | 0.036 | 0.032 | 0.255 | 0.801 | 0.084 | 0.046 | 0.065 |
| rs9888533 | T | C | 0.508 | -0.028 | 0.023 | 0.220 | 0.529 | -0.041 | 0.034 | 0.233 |
| rs9926784 | T | C | 0.759 | -0.006 | 0.029 | 0.830 | 0.764 | -0.048 | 0.042 | 0.256 |
| rs9944219 | A | G | 0.611 | 0.002 | 0.023 | 0.934 | 0.607 | 0.019 | 0.034 | 0.564 |
| rs994596^a^ | T | C | 0.325 | -0.053 | 0.024 | 0.026 |  |  |  |  |
| rs9951619 | T | G | 0.277 | -0.037 | 0.026 | 0.152 | 0.299 | -0.085 | 0.039 | 0.027 |
| rs10197031^b^ | T | C |  |  |  |  | 0.677 | 0.075 | 0.037 | 0.043 |
| rs10510419^b^ | T | G |  |  |  |  | 0.192 | -0.083 | 0.046 | 0.071 |
| rs11611246 ^b^ | T | G |  |  |  |  | 0.229 | 0.084 | 0.042 | 0.048 |
| rs429358 ^b^ | T | C |  |  |  |  | 0.817 | -0.057 | 0.050 | 0.254 |
| rs57989773 ^b^ | T | C |  |  |  |  | 0.742 | 0.085 | 0.040 | 0.031 |

Abbrevations: SNP, Single Nucleotide Polymorphism; EA, effect allele; OA, other allele; EAF, effect allele frequency; SE, standard error.

^a^ Heterogeneous SNP identified as outliers by the radial regression in case of COVID-19 hospitalization.

^b^ Heterogeneous SNP identified as outliers by the radial regression in case of COVID-19 susceptibility.

**Supplementary Table S2** Associations of genome-wide significant SNPs used as instruments for waist circumference and the susceptibility as well as hospitalization due to COVID-19

|  |  |  | **COVID-19 susceptibility (n= 1079768)** | | | | **COVID-19 hospitalization (n=900687)** | | | |
| --- | --- | --- | --- | --- | --- | --- | --- | --- | --- | --- |
| **SNP** | **EA** | **OA** | **EAF** | $\boldsymbol{\beta}$ | **SE** | **P** | **EAF** | $\boldsymbol{\beta}$ | **SE** | **P** |
| rs10132280 | A | C | 0.334 | 0.016 | 0.024 | 0.490 | 0.332 | 0.047 | 0.035 | 0.181 |
| rs10767658 | G | C | 0.693 | -0.032 | 0.024 | 0.186 | 0.678 | -0.007 | 0.037 | 0.850 |
| rs10840100 | G | A | 0.599 | 0.001 | 0.023 | 0.949 | 0.597 | -0.020 | 0.034 | 0.562 |
| rs10938397 | G | A | 0.408 | -0.007 | 0.022 | 0.749 | 0.430 | -0.015 | 0.033 | 0.661 |
| rs10968576 | G | A | 0.281 | -0.003 | 0.025 | 0.910 | 0.305 | 0.028 | 0.037 | 0.446 |
| rs11165623 | A | G | 0.448 | 0.006 | 0.022 | 0.782 | 0.464 | 0.020 | 0.033 | 0.534 |
| rs12429545 | A | G | 0.158 | 0.046 | 0.033 | 0.162 | 0.182 | 0.025 | 0.049 | 0.611 |
| rs1516725 | C | T | 0.836 | 0.013 | 0.032 | 0.685 | 0.817 | -0.010 | 0.047 | 0.837 |
| rs1558902 | A | T | 0.386 | 0.012 | 0.023 | 0.591 | 0.407 | 0.055 | 0.033 | 0.097 |
| rs16894959 | C | T | 0.164 | 0.032 | 0.032 | 0.325 | 0.179 | 0.052 | 0.048 | 0.276 |
| rs16996700 | C | T | 0.279 | 0.001 | 0.025 | 0.971 | 0.297 | 0.018 | 0.036 | 0.617 |
| rs17066856 | C | T | 0.164 | -0.028 | 0.038 | 0.456 | 0.172 | -0.045 | 0.055 | 0.418 |
| rs17381664 | C | T | 0.335 | 0.053 | 0.024 | 0.027 | 0.362 | 0.073 | 0.036 | 0.043 |
| rs2033529 | G | A | 0.269 | 0.020 | 0.029 | 0.498 | 0.312 | 0.041 | 0.038 | 0.287 |
| rs2112347 | G | T | 0.412 | 0.040 | 0.023 | 0.079 | 0.411 | 0.062 | 0.034 | 0.068 |
| rs2287019 | T | C | 0.199 | -0.001 | 0.028 | 0.980 | 0.223 | -0.073 | 0.043 | 0.088 |
| rs2293576 | A | G | 0.329 | -0.027 | 0.024 | 0.248 | 0.356 | -0.013 | 0.035 | 0.712 |
| rs2325036 | C | A | 0.403 | -0.005 | 0.023 | 0.831 | 0.416 | 0.001 | 0.034 | 0.965 |
| rs2489623 | C | A | 0.529 | -0.033 | 0.022 | 0.137 | 0.524 | -0.011 | 0.033 | 0.734 |
| rs2531992 | G | A | 0.774 | 0.020 | 0.031 | 0.521 | 0.766 | -0.018 | 0.046 | 0.688 |
| rs2820292 | C | A | 0.526 | 0.032 | 0.022 | 0.144 | 0.534 | 0.037 | 0.033 | 0.260 |
| rs3127553 | A | G | 0.615 | -0.021 | 0.023 | 0.352 | 0.600 | -0.017 | 0.034 | 0.619 |
| rs3849570 | A | C | 0.367 | 0.004 | 0.023 | 0.852 | 0.373 | 0.043 | 0.035 | 0.220 |
| rs4776970 | T | A | 0.417 | -0.010 | 0.023 | 0.666 | 0.403 | -0.040 | 0.034 | 0.241 |
| rs6163 | A | C | 0.396 | 0.045 | 0.023 | 0.056 | 0.411 | 0.048 | 0.034 | 0.164 |
| rs633715 | C | T | 0.198 | 0.003 | 0.028 | 0.920 | 0.222 | 0.054 | 0.043 | 0.207 |
| rs6440003 | A | G | 0.453 | 0.006 | 0.023 | 0.805 | 0.426 | -0.059 | 0.034 | 0.080 |
| rs6545714 | A | G | 0.622 | -0.020 | 0.023 | 0.373 | 0.615 | -0.035 | 0.034 | 0.301 |
| rs6567160 | C | T | 0.265 | 0.015 | 0.029 | 0.593 | 0.275 | -0.004 | 0.039 | 0.924 |
| rs6755502 | C | T | 0.815 | 0.034 | 0.030 | 0.260 | 0.789 | -0.006 | 0.043 | 0.896 |
| rs7138803 | A | G | 0.364 | 0.015 | 0.023 | 0.512 | 0.376 | 0.028 | 0.034 | 0.414 |
| rs7144011 | T | G | 0.212 | 0.003 | 0.028 | 0.915 | 0.230 | 0.015 | 0.041 | 0.711 |
| rs7239883 | A | G | 0.612 | 0.005 | 0.023 | 0.815 | 0.610 | -0.003 | 0.034 | 0.928 |
| rs749671 | A | G | 0.349 | 0.016 | 0.023 | 0.480 | 0.380 | 0.028 | 0.033 | 0.406 |
| rs7498665 | G | A | 0.348 | -0.003 | 0.023 | 0.913 | 0.364 | 0.013 | 0.035 | 0.700 |
| rs7531118 | C | T | 0.484 | -0.026 | 0.022 | 0.247 | 0.532 | -0.038 | 0.034 | 0.259 |
| rs7550711 | T | C | 0.052 | 0.124 | 0.067 | 0.063 | 0.085 | 0.189 | 0.111 | 0.088 |
| rs7903146 | T | C | 0.312 | 0.023 | 0.024 | 0.332 | 0.325 | 0.043 | 0.035 | 0.226 |
| rs806794 | G | A | 0.338 | 0.013 | 0.024 | 0.588 | 0.364 | 0.006 | 0.036 | 0.864 |
| rs929641 | G | A | 0.429 | -0.008 | 0.022 | 0.717 | 0.433 | -0.037 | 0.033 | 0.267 |
| rs9400239 | C | T | 0.615 | 0.030 | 0.024 | 0.204 | 0.619 | 0.065 | 0.035 | 0.062 |
| rs943005 | T | C | 0.189 | 0.022 | 0.029 | 0.446 | 0.218 | 0.020 | 0.044 | 0.657 |

Abbrevations: SNP, Single Nucleotide Polymorphism; EA, effect allele; OA, other allele; EAF, effect allele frequency; SE, standard error.

**Supplementary Table S3** Associations of genome-wide significant SNPs used as instruments for trunk fat ratio and the susceptibility as well as hospitalization due to COVID-19

| **SNP** | **EA** | **OA** | **EAF** | $\boldsymbol{\beta}$ | **SE** | **P** | **EAF** | $\boldsymbol{\beta}$ | **SE** | **P** |
| --- | --- | --- | --- | --- | --- | --- | --- | --- | --- | --- |
| rs10402308 | A | G | 0.248 | -0.047 | 0.028 | 0.096 | 0.253 | -0.065 | 0.043 | 0.133 |
| rs10962638 | A | G | 0.152 | -0.044 | 0.032 | 0.166 | 0.172 | -0.030 | 0.048 | 0.532 |
| rs11049533 | G | A | 0.244 | -0.024 | 0.025 | 0.337 | 0.271 | -0.037 | 0.037 | 0.325 |
| rs11205303 | C | T | 0.372 | 0.015 | 0.022 | 0.500 | 0.386 | 0.006 | 0.033 | 0.850 |
| rs115912456 | G | A | 0.072 | -0.123 | 0.056 | 0.027 | 0.107 | -0.174 | 0.083 | 0.037 |
| rs12790261 | A | C | 0.091 | -0.046 | 0.047 | 0.323 | 0.142 | -0.070 | 0.069 | 0.312 |
| rs12905253 | A | G | 0.428 | -0.014 | 0.022 | 0.528 | 0.442 | -0.025 | 0.033 | 0.445 |
| rs143384 | G | A | 0.497 | 0.016 | 0.023 | 0.472 | 0.463 | 0.017 | 0.033 | 0.612 |
| rs17511102 | T | A | 0.104 | 0.010 | 0.041 | 0.814 | 0.152 | 0.057 | 0.064 | 0.375 |
| rs1986599 | G | T | 0.113 | 0.027 | 0.037 | 0.465 | 0.142 | 0.051 | 0.052 | 0.324 |
| rs2071167 | T | C | 0.295 | 0.026 | 0.026 | 0.319 | 0.296 | 0.006 | 0.038 | 0.875 |
| rs2274432 | A | G | 0.338 | 0.027 | 0.023 | 0.240 | 0.356 | 0.037 | 0.035 | 0.296 |
| rs314263 | C | T | 0.322 | 0.012 | 0.024 | 0.614 | 0.328 | 0.054 | 0.036 | 0.131 |
| rs35650604 | G | A | 0.142 | 0.020 | 0.034 | 0.551 | 0.188 | -0.003 | 0.048 | 0.954 |
| rs3791679 | G | A | 0.245 | -0.030 | 0.026 | 0.254 | 0.267 | -0.025 | 0.038 | 0.514 |
| rs3817428 | G | C | 0.235 | 0.045 | 0.026 | 0.083 | 0.265 | 0.045 | 0.039 | 0.251 |
| rs41271299 | T | C | 0.072 | 0.047 | 0.057 | 0.408 | 0.121 | 0.066 | 0.082 | 0.417 |
| rs4483821 | G | A | 0.531 | 0.008 | 0.025 | 0.763 | 0.511 | 0.026 | 0.033 | 0.433 |
| rs459193 | A | G | 0.316 | 0.019 | 0.025 | 0.446 | 0.320 | -0.015 | 0.037 | 0.689 |
| rs4694510 | A | C | 0.095 | 0.038 | 0.046 | 0.406 | 0.120 | 0.156 | 0.070 | 0.025 |
| rs4846204 | T | C | 0.148 | 0.007 | 0.032 | 0.830 | 0.171 | 0.051 | 0.049 | 0.303 |
| rs6785012 | T | C | 0.448 | 0.005 | 0.023 | 0.814 | 0.420 | -0.059 | 0.034 | 0.084 |
| rs7236575 | A | G | 0.189 | 0.013 | 0.031 | 0.672 | 0.205 | -0.008 | 0.046 | 0.866 |
| rs72708236 | G | T | 0.055 | -0.067 | 0.064 | 0.296 | 0.094 | -0.140 | 0.101 | 0.167 |
| rs72755233 | A | G | 0.116 | 0.021 | 0.038 | 0.582 | 0.175 | 0.035 | 0.061 | 0.564 |
| rs7680661 | G | A | 0.199 | 0.013 | 0.029 | 0.648 | 0.219 | 0.005 | 0.042 | 0.913 |
| rs77485628 | T | C | 0.131 | 0.010 | 0.035 | 0.768 | 0.159 | -0.057 | 0.051 | 0.267 |
| rs7763064 | A | G | 0.338 | -0.015 | 0.024 | 0.532 | 0.335 | -0.059 | 0.035 | 0.092 |
| rs79112217 | C | T | 0.106 | 0.017 | 0.037 | 0.653 | 0.132 | -0.047 | 0.055 | 0.393 |
| rs798491 | G | A | 0.263 | -0.004 | 0.025 | 0.860 | 0.302 | -0.022 | 0.036 | 0.543 |
| rs888762 | C | A | 0.293 | -0.004 | 0.024 | 0.881 | 0.320 | 0.014 | 0.036 | 0.707 |
| rs9393688 | T | A | 0.302 | 0.014 | 0.025 | 0.578 | 0.355 | 0.006 | 0.038 | 0.881 |
| rs75848127^a^ | A | G |  |  |  |  | 0.207 | -0.087 | 0.047 | 0.066 |

Abbrevations: SNP, Single Nucleotide Polymorphism; EA, effect allele; OA, other allele; EAF, effect allele frequency; SE, standard error.

^a^ Heterogeneous SNP identified as outliers by the radial regression in case of COVID-19 susceptibility.

**Supplementary Table S4** Power analysis for the associations between genetically predicted BMI, WC and TFR (continuous) and COVID-19 susceptibility as well as hospitalization (binary)

| **Exposure** | **Outcome** | **N cases** | **N controls** | **OR=1.1** | **OR=1.2** | **OR=1.3** | **OR=1.4** | **OR=1.5** | **OR=1.6** | **OR=1.7** | **OR=1.8** |
| --- | --- | --- | --- | --- | --- | --- | --- | --- | --- | --- | --- |
| BMI | Suscept. | 6696 | 1073072 | 0.477 | 0.967 | 1.000 | 1.000 | 1.000 | 1.000 | 1.000 | 1.000 |
| WC | Suscept. | 6696 | 1073072 | 0.136 | 0.397 | 0.721 | 0.924 | 0.989 | 0.999 | 1.000 | 1.000 |
| TFR | Suscept. | 6696 | 1073072 | 0.147 | 0.437 | 0.771 | 0.950 | 0.994 | 1.000 | 1.000 | 1.000 |
| BMI | Hosp. | 3199 | 897488 | 0.262 | 0.752 | 0.977 | 1.000 | 1.000 | 1.000 | 1.000 | 1.000 |
| WC | Hosp. | 3199 | 897488 | 0.090 | 0.217 | 0.422 | 0.652 | 0.836 | 0.941 | 0.984 | 0.997 |
| TFR | Hosp. | 3199 | 897488 | 0.097 | 0.243 | 0.473 | 0.713 | 0.883 | 0.966 | 0.993 | 0.999 |

Abbrevations: OR, odds ratio; BMI, body mass index; WC, waist circumference; TFR, trunk fat ratio.

**Supplementary Table S5** Results of the MR-PRESSO global and Egger-intercept tests for detecting horizontal and directional pleiotropy in the univariable Mendelian randomization setting

| **Exposure** | **Outcome** | **MR-PRESSO RSSobs** | **P_RSS_** | **Egger-intercept** | **SE** | **P_Egger_** |
| --- | --- | --- | --- | --- | --- | --- |
| BMI | susceptibility | 499.022 | 0.726 | 0.087 | 0.123 | 0.480 |
| WC | susceptibility | 33.796 | 0.840 | 0.377 | 0.391 | 0.340 |
| TFR | susceptibility | 21.365 | 0.937 | 0.516 | 0.776 | 0.511 |
| BMI | hospitalization | 522.938 | 0.459 | 0.084 | 0.125 | 0.504 |
| WC | hospitalization | 40.897 | 0.556 | 0.036 | 0.427 | 0.934 |
| TFR | hospitalization | 35.942 | 0.368 | 0.304 | 1.014 | 0.766 |

Abbrevations: MR-PRESSO, Mendelian Randomization Pleiotropy RESidual Sum and Outlier; RSSobs, observed residual sum of squares; SE, standard error; BMI, body mass index; WC, waist circumference; TFR, trunk fat ratio.

**Supplementary Table S6** Between SNP-heterogeneity based on the radial regression framework in the univariable Mendelian randomization setting

| **Exposure** | **Outcome** | **Cochran‘s Q** | **df** | **P_Q_** | **Rueckers Q'** | **P_Q'_** | **Q-Q'** | **P_Q-Q'_** | **Q'/Q** |
| --- | --- | --- | --- | --- | --- | --- | --- | --- | --- |
| BMI | susceptibility | 508.480 | 535 | 0.789 | 508.147 | 0.792 | 0.333 | 0.564 | 0.999 |
| WC | susceptibility | 32.492 | 41 | 0.826 | 31.815 | 0.848 | 0.677 | 0.411 | 0.979 |
| TFR | susceptibility | 19.907 | 32 | 0.953 | 19.706 | 0.956 | 0.201 | 0.654 | 0.990 |
| BMI | hospitalization | 532.830 | 535 | 0.518 | 532.583 | 0.521 | 0.247 | 0.619 | 1.000 |
| WC | hospitalization | 39.176 | 41 | 0.552 | 39.184 | 0.552 | -0.008 | 1.000 | 1.000 |
| TFR | hospitalization | 34.261 | 33 | 0.407 | 34.258 | 0.407 | 0.003 | 0.956 | 1.000 |

Abbrevations: BMI, body mass index; WC, waist circumference; TFR, trunk fat ratio; df, degrees of freedom.

**Supplementary Table S7** Influential Single Nucleotide Polymorphisms (SNPs) identified and excluded by an iterative approach calculating SNP-specific Q_j_-statistics in the radial inverse-variance weighted as well as MR-Egger methods for the impact of body composition measures on COVID-19 susceptibility and severity

| **SNP** | $\boldsymbol{Q}_{\boldsymbol{IVW}}$ | $\boldsymbol{P}_{\boldsymbol{IVW}}$ | $\boldsymbol{Q}_{\boldsymbol{Egger}}$ | $\boldsymbol{P}_{\boldsymbol{Egger}}$ | **Iteration** |
| --- | --- | --- | --- | --- | --- |
| **Body Mass Index on COVID-19 susceptibility** | | | | | |
| rs10197031 | 9.182 | 0.002 | 9.194 | 0.002 | 1 |
| rs10510419 | 8.15 | 0.004 | 8.08 | 0.004 | 1 |
| rs429358 | 7.631 | 0.006 | 7.553 | 0.006 | 1 |
| rs11611246 | 7.387 | 0.007 | 7.458 | 0.006 | 1 |
| rs57989773 | 6.926 | 0.008 | 7.003 | 0.008 | 1 |
| **Body Mass Index on COVID-19 hospitalization** | | | | | |
| rs994596 | 8.983 | 0.003 | 9.148 | 0.002 | 1 |
| rs4988235 | 8.041 | 0.005 | 8.32 | 0.004 | 1 |
| rs4655141 | 7.445 | 0.006 | 7.516 | 0.006 | 1 |
| rs12681792 | 7.058 | 0.008 | 7.249 | 0.007 | 1 |
| rs9370410 | 6.669 | 0.010 |  |  | 2 |
| **Trunk fat ratio** **on COVID-19 susceptibility** | | | | | |
| rs75848127 | 9.217 | 0.002 | 9.205 | 0.002 | 1 |

Abbrevations: SNP, Single Nucleotide Polymorphisms.

**Supplementary Table S8** Results of Egger-intercept tests for detecting directional pleiotropy in the multivariable Mendelian randomization approach

|  | **Susceptibility** | | | **Hospitalization** | | |
| --- | --- | --- | --- | --- | --- | --- |
| **Exposures** | **Egger-intercept** | **SE** | **P** | **Egger-intercept** | **SE** | **P** |
| BMI, WC | 0.001 | 0.003 | 0.875 | 0.005 | 0.005 | 0.300 |
| BMI, TFR | -0.001 | 0.004 | 0.748 | 0.012 | 0.006 | 0.044 |

Abbrevations: BMI, body mass index; WC, waist circumference; TFR, trunk fat ratio; SE, standard error.

**Supplementary Table S9** Heterogeneity statistics in the multivariable Mendelian randomization approach.

|  |  | **Susceptibility** | | **Hospitalization** | |
| --- | --- | --- | --- | --- | --- |
| **Method** | **Exposures** | **Q-statistic** | **P** | **Q-statistic** | **P** |
| Robust IVW (mult. rand. effects) | BMI, WC | 523.238 | 0.079 | 527.924 | 0.057 |
| MR-Egger (rand. effects) | BMI, WC | 482.977 | 0.428 | 517.851 | 0.095 |
| Robust IVW (mult. rand. effects) | BMI, TFR | 289.130 | 0.112 | 256.222 | 0.572 |
| MR-Egger (rand. effects) | BMI, TFR | 254.452 | 0.585 | 251.090 | 0.643 |

Abbrevations: BMI, body mass index; WC, waist circumference; TFR, trunk fat ratio; IVW (mult. rand. effects), inverse-variance weighted model with multiplicative random effects.

**Supplementary Table S10** Results of Egger-intercept tests for detecting directional pleiotropy within the multivariable Mendelian randomization mediation analysis.

|  |  | **Susceptibility** | | | **Hospitalization** | | |
| --- | --- | --- | --- | --- | --- | --- | --- |
| **Exposure** | **Mediators** | **Egger-intercept** | **SE** | **P** | **Egger-intercept** | **SE** | **P** |
| BMI | T2D | 0.001 | 0.003 | 0.709 | 0.006 | 0.005 | 0.246 |
| WC | T2D | -0.008 | 0.007 | 0.229 | -0.002 | 0.011 | 0.867 |
| TFR | T2D | -0.008 | 0.017 | 0.644 | -0.015 | 0.027 | 0.582 |
| BMI | CVD | 0.000 | 0.003 | 0.902 | 0.004 | 0.005 | 0.497 |
| WC | CVD | -0.003 | 0.005 | 0.594 | -0.005 | 0.009 | 0.597 |
| TFR | CVD | 0.006 | 0.013 | 0.663 | -0.009 | 0.019 | 0.630 |
| BMI | T2D, CVD | 0.002 | 0.004 | 0.675 | 0.005 | 0.006 | 0.346 |
| WC | T2D, CVD | -0.001 | 0.006 | 0.865 | -0.006 | 0.010 | 0.540 |
| TFR | T2D, CVD | 0.003 | 0.014 | 0.825 | -0.014 | 0.021 | 0.505 |

Abbrevations: BMI, body mass index; WC, waist circumference; TFR, trunk fat ratio; T2D, type 2 diabetes; CVD cardio vascular diseases; SE, standard error.

**Supplementary Table S11** Heterogeneity statistics from the multivariable Mendelian randomization mediation analyses.

|  |  |  | **Susceptibility** | | **Hospitalization** | |
| --- | --- | --- | --- | --- | --- | --- |
| **Method** | **Exposure** | **Mediators** | **Q-statistic** | **P** | **Q-statistic** | **P** |
| Robust IVW (mult. rand. effects) | BMI | T2D | 515.084 | 0.008 | 517.972 | 0.006 |
| MR-Egger (rand. effects) | BMI | T2D | 462.797 | 0.218 | 484.487 | 0.066 |
| Robust IVW (mult. rand. effects) | WC | T2D | 31.049 | 0.659 | 36.690 | 0.390 |
| MR-Egger (rand. effects) | WC | T2D | 32.406 | 0.546 | 39.751 | 0.229 |
| Robust IVW (mult. rand. effects) | TFR | T2D | 10.809 | 0.930 | 9.526 | 0.964 |
| MR-Egger (rand. effects) | TFR | T2D | 18.319 | 0.435 | 20.048 | 0.330 |
| Robust IVW (mult. rand. effects) | BMI | CVD | 469.247 | 0.111 | 465.256 | 0.130 |
| MR-Egger (rand. effects) | BMI | CVD | 428.787 | 0.535 | 461.751 | 0.148 |
| Robust IVW (mult. rand. effects) | WC | CVD | 54.436 | 0.345 | 67.466 | 0.061 |
| MR-Egger (rand. effects) | WC | CVD | 54.456 | 0.309 | 67.687 | 0.048 |
| Robust IVW (mult. rand. effects) | TFR | CVD | 4.843 | 0.978 | 9.407 | 0.742 |
| MR-Egger (rand. effects) | TFR | CVD | 8.561 | 0.740 | 7.137 | 0.848 |
| Robust IVW (mult. rand. effects) | BMI | T2D, CVD | 422.197 | 0.019 | 426.110 | 0.012 |
| MR-Egger (rand. effects) | BMI | T2D, CVD | 381.032 | 0.247 | 408.733 | 0.045 |
| Robust IVW (mult. rand. effects) | WC | T2D, CVD | 40.686 | 0.440 | 43.327 | 0.331 |
| MR-Egger (rand. effects) | WC | T2D, CVD | 50.877 | 0.096 | 63.560 | 0.008 |
| Robust IVW (mult. rand. effects) | TFR | T2D, CVD | 4.291 | 0.830 | 3.688 | 0.884 |
| MR-Egger (rand. effects) | TFR | T2D, CVD | 2.694 | 0.912 | 5.287 | 0.625 |

Abbrevations: BMI, body mass index; WC, waist circumference; TFR, trunk fat ratio; T2D, type 2 diabetes; CVD cardio vascular diseases; IVW (mult. rand. effects), inverse-variance weighted model with multiplicative random effects.

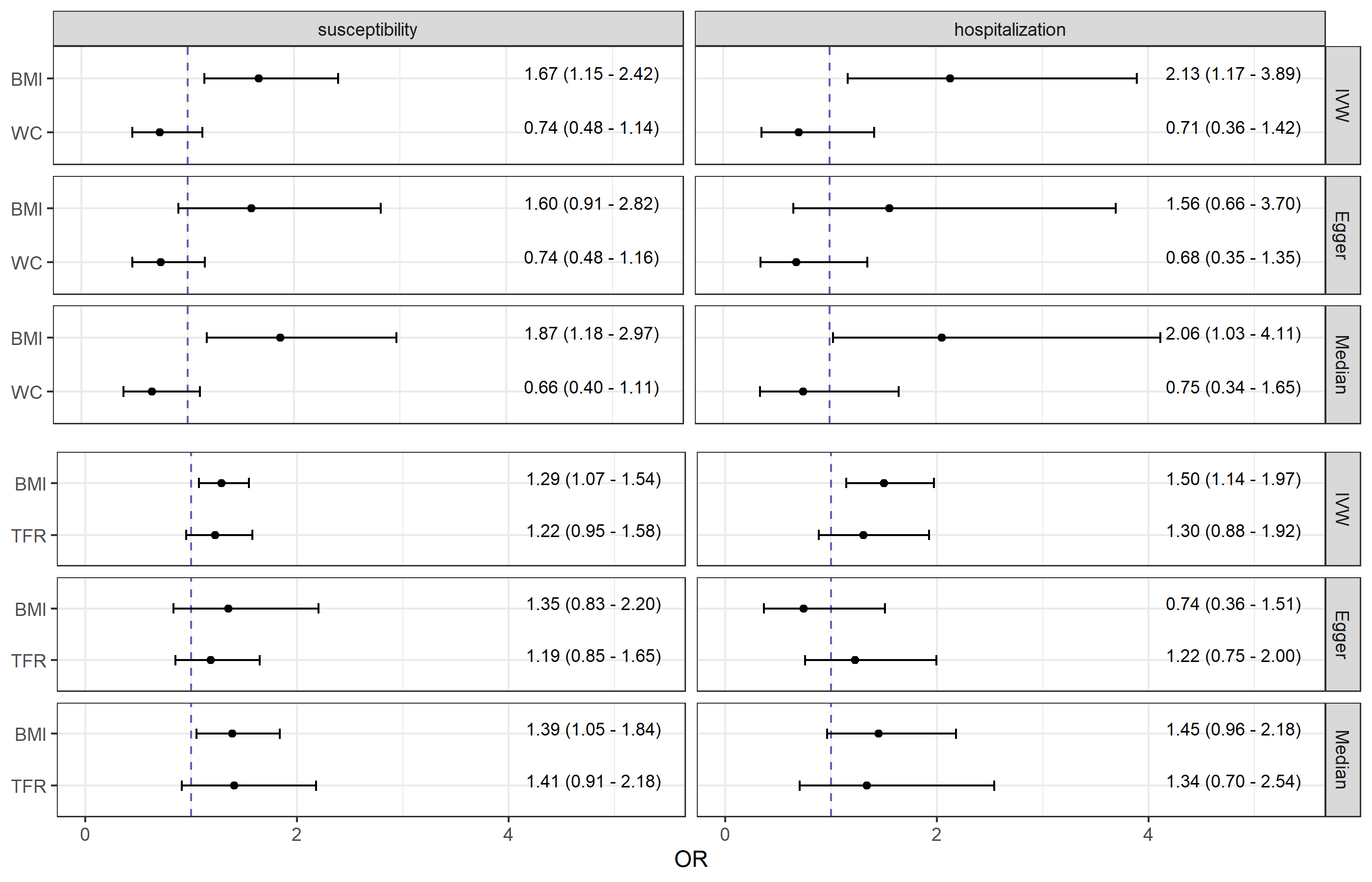

**Supplementary Figure S12** Causal direct effect estimates (odds ratios and 95 % confidence intervals) from pairwise multivariable Mendelian randomization analyses of body mass index (BMI), waist circumference (WC), and trunk fat ratio (TFR) with COVID-19 susceptibility as well as hospitalization. In main analysis the robust inverse-variance weighted (IVW) method with multiplicative random effects was applied. In sensitivity analyses the MR-Egger with multiplicative random effects and the Median approach were performed.

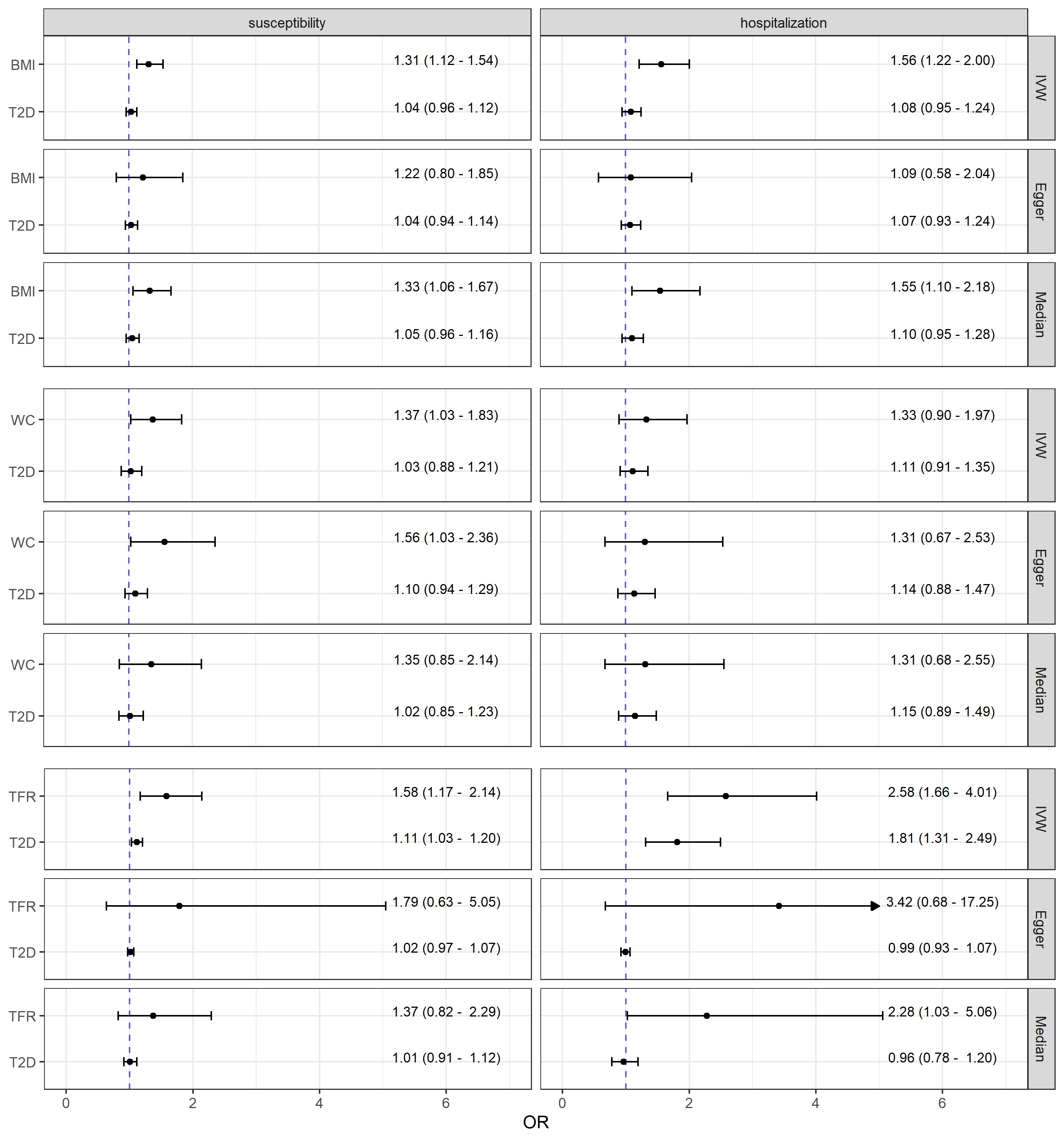

**Supplementary Figure S13** Causal estimates (odds ratios and 95 % confidence intervals) from multivariable Mendelian randomization mediation analyses of body composition measures, body mass index (BMI), waist circumference (WC), and trunk fat ratio (TFR), adjusted for type 2 diabetes (T2D) on COVID-19 susceptibility as well as hospitalization. In main analysis the robust inverse-variance weighted (IVW) method with multiplicative random effects was applied. In sensitivity analyses the MR-Egger with multiplicative random effects and the Median approach were performed. The arrow represents a confidence interval exceeding the plot range (Upper CI = 17.25).

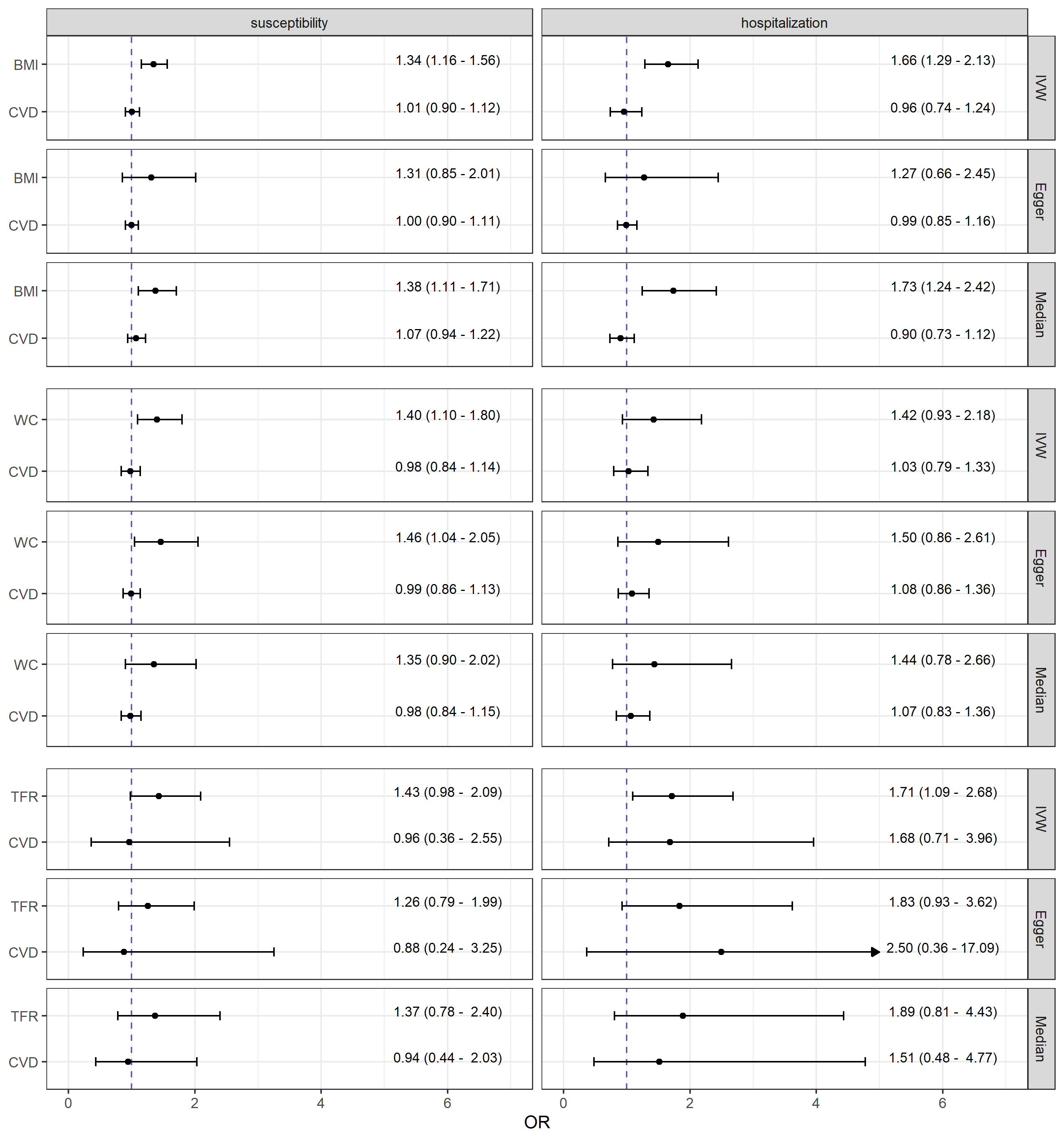

**Supplementary Figure S14** Causal estimates (odds ratios and 95 % confidence intervals) from multivariable Mendelian randomization mediation analyses of body composition measures, body mass index (BMI), waist circumference (WC), and trunk fat ratio (TFR), adjusted for cardiovascular diseases (CVD) on COVID-19 susceptibility as well as hospitalization. In main analysis the robust inverse-variance weighted (IVW) method with multiplicative random effects was applied. In sensitivity analyses the MR-Egger with multiplicative random effects and the Median approach were performed. The arrow represents a confidence interval exceeding the plot range (Upper CI = 17.09).

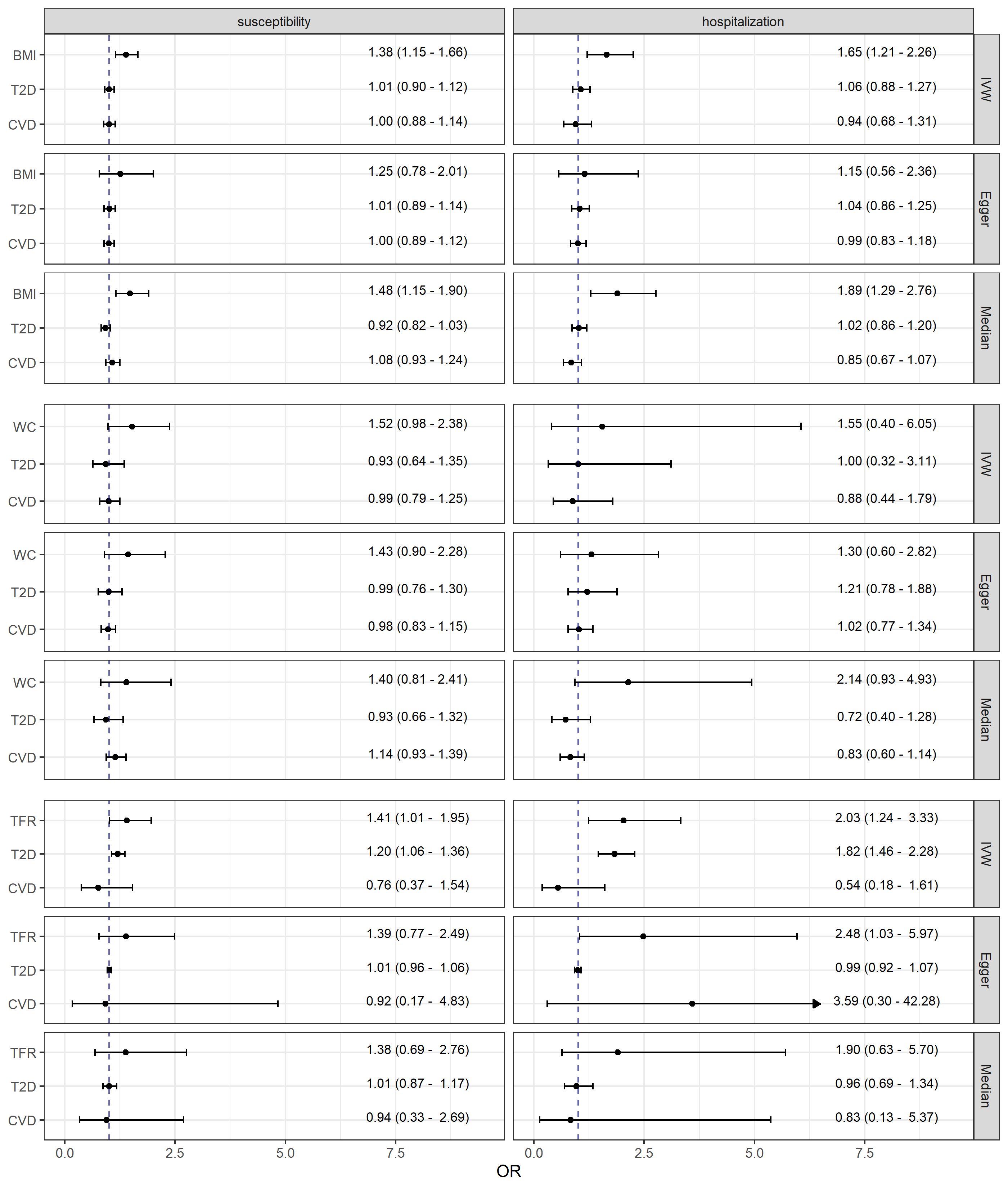

**Supplementary Figure S15** Causal estimates (odds ratios and 95 % confidence intervals) from multivariable Mendelian randomization mediation analyses of body composition measures, body mass index (BMI), waist circumference (WC), and trunk fat ratio (TFR), adjusted for both, type 2 diabetes (T2D) and cardiovascular diseases (CVD) on COVID-19 susceptibility as well as hospitalization. In main analysis the robust inverse-variance weighted (IVW) method with multiplicative random effects was applied. In sensitivity analyses the MR-Egger with multiplicative random effects and the Median approach were performed. The arrow represents a confidence interval exceeding the plot range (Upper CI = 42.28).
